# Sex-specific omics scores of sex hormones and associations with metabolic and sleep disorders

**DOI:** 10.64898/2026.04.27.26351843

**Authors:** Ziqing Wang, Yu Zhang, Alexis C Wood, Mark D Benson, Kent D Taylor, Xiuqing Guo, Stephen S Rich, Robert E Gerszten, Susan Redline, Jerome I. Rotter, Peter Ganz, Rajat Deo, Ruth Dubin, Peter Y. Liu, Tamar Sofer

**Author notes:** Corresponding author: Ziqing Wang.

## Abstract

**Introduction:** Sex hormones shape biological sex differences and alter the onset and severity of sleep and metabolic diseases in a sex-specific manner. To better understand relationships and underlying mechanisms, we develop summary proteomics and metabolomics scores for sex hormones and investigate their associations with sleep and metabolic disorders.

**Methods:** We used proteome-(n= 3680) and metabolome-wide (n= 1649) data from the baseline exam of the Multi-Ethnic Study of Atherosclerosis (MESA) cohort to develop female- and male-specific omics scores for sex hormones including total (Total T), bioavailable (Bio T), and free (Free T) testosterone, estradiol (E2) and sex hormone binding protein (SHBG). Each omics dataset was randomly split assigning 80% of participants to a training dataset and the remaining 20% to a test dataset. We applied linear regression with bootstrap standard errors, adjusting for age, BMI, self-reported race/ethnicity and study site, to identify sex hormone-associated proteins and metabolites (i.e FDR< .05). Lasso penalized regression was then used to select independent features, from which weighted protein (ProtS) and metabolite scores (MetS) were constructed as weighted sums, and examined in the validation dataset. Subsequently, we conducted sex-stratified association analysis of the validated omics scores using data from MESA baseline, exams 4 (proteomics) and 5 (proteomics, metabolomics) with sleep and metabolic phenotypes, timepoints where sex hormones were not measured.

**Results:** All constructed omics scores were significantly associated with their corresponding hormones in the test dataset. Higher omics scores of SHBG and lower omics scores of Free T were associated with lower diabetes risk in both sexes; and higher E2 scores with higher incident hypertension risk only in men. In males, Total T had protective diabetes associations, whereas in females they were linked to greater risk. Similarly, higher ProtS-Free T and lower ProtS-SHBG were associated with increased risk for OSA in both sexes. Finally, higher E2 scores were associated with higher risk of insomnia only in males.

**Conclusions:** Summary omics-based scores reveal sex-specific cross-sectional associations with sleep and incident metabolic disorders. These findings highlight the potential of these omics proxies to improve risk stratification and generate insights into mechanisms linking sex hormones with disease.

## Introduction

Sex hormones play a critical role in sexual phenotypic differentiation and sex differences in disease susceptibilities ^1,2^. For instance, men with higher total testosterone (Total T) levels demonstrate a lower risk of metabolic syndrome, while women with higher Total T levels show increased risks^3^. Higher genetically determined Total T increases the risk of diabetes in men, whereas genetically determined bioavailable testosterone (Bio T) decreases it in women^4^. Higher levels of estradiol (E2) and lower levels of sex binding hormones (SHBG) are associated with increased risk of incident diabetes in both men and women^5^. In addition to cardiovascular and metabolic diseases, sleep disorders also manifest strong sex differences due to complex relationships among sleep patterns, hormonal levels and environmental factors. Two prevalent sleep disorders with pronounced sex differences are obstructive sleep apnea (OSA) and insomnia.

OSA prevalence is higher in men, and is characterized by sex-specific pathophysiological endotypes, with women demonstrating lower upper-airway collapsibility and lower arousal thresholds than men^6^. Such OSA-related traits in females become more prevalent across the menopause transition, with distinct clinical symptoms between pre- and postmenopausal women^7^ that improve with hormone replacement therapy^8^. In parallel to hormonal changes, women undergoing hormone therapy exhibit more stable breathing and lower apneic thresholds compared to those without hormone therapy and to males^9^. In contrast, insomnia is more prevalent in females than in males with differences first seen around the time of puberty and then persist across the lifespan^10^. These differences are considered to partly reflect the effects of sex hormones on sleep architecture and circadian hormone release^11,12^.

While specific disease mechanisms remain to be elucidated, sex hormones may influence disease severity and onset by regulating various physiological processes, including oxidative stress, immune response, body fat distribution, upper airway anatomy, and insulin resistance^7,13^. For example, sex-dependent patterns of fat distribution are largely modulated by sex hormones^14^, and hence, tend to attenuate after menopause^10^. Relevant to OSA, the longer upper airway observed in males becomes evident after puberty, and is associated with increased airway collapsibility^15,16^. Therefore, understanding the underlying mechanisms behind sex hormone modulation is essential to reduce clinical bias and to advance potentially sex-specific therapeutic strategies and personalized medicine.

High-throughput omics data, such as genomics, epigenetics, metabolomics, and proteomics, have advanced our understanding of unique molecular profiles specific to particular traits, leading to more accurate population risk stratification through the construction of omics-based proxies. In alignment with the measured hormone levels, higher genetically-predicted SHBG levels were also associated with reduced risk of hypertension, type 2 diabetes and coronary atherosclerotic outcomes^17^. While increasing research has focused on sex differences in disease pathology^7,16–19^, cardiovascular, sleep and cognitive outcomes, as well as changes in sex hormone levels reflect complex and intricate relationships determined by both biological sex and sociocultural factors^20^. As omics data have become increasingly available, this study aims to develop proteomics and metabolomics scores that serve as proxy variables for sex hormone levels. We investigate the application of such omics scores to study the relationship between biological sex and sex-biased metabolic and sleep disorders, and to investigate the omics signatures of sex hormones to explore underlying biological mechanisms.

## Methods

### The Multi-Ethnic Study of Atherosclerosis (MESA) study

MESA is a multi-ethnic longitudinal community-based study following 6,814 participants aged 45–84 years at baseline from six field centers (Baltimore, MD; Chicago, IL; Forsyth County, NC; Los Angeles, CA; New York, NY; and St Paul, MN)^21^. At baseline (Exam 1; 2000-2002), all participants were free of overt clinical cardiovascular diseases, and provided self-reported information on sociodemographic characteristics, including age, sex, race (White, African American, Hispanic, and Chinese American), and menopausal status (postmenopausal: yes or no), using standard questionnaires^21^. Diabetes was defined by fasting plasma glucose ≥126 mg/dL and/or use of insulin or other diabetes medications^22^. Hypertension was characterized as systolic blood pressure ≥ 140 mmHg, diastolic blood pressure ≥ 90 mmHg, use of antihypertensive medication, and/or self-reported prior diagnosis^22^.

### Hormone profiling

Serum hormone concentrations were measured from an early morning fasting blood sample at baseline and processed at the University of Massachusetts Medical Center in Worcester, MA^23^. Sex hormones were not measured at other exams. Total testosterone (Total T) and sex hormone binding globulin (SHBG) were measured using radioimmunoassay and chemiluminescent enzyme immunometric assay (Diagnostic Products Corporation, Los Angeles, CA), respectively^23^. E2 was measured using an ultrasensitive radioimmunoassay kit (Diagnostic System Laboratories, Webster, TX)^23^. Hormone samples were checked for assay variability using 5% blind quality control samples from each batch^23^. Free testosterone (Free T) was calculated as a percentage of Total T, [total T × (percent-Free T × 0.01)]^24^, and Bio T was computed from Total T and SHBG concentrations^25^. A total of 5710 participants with hormone data remained after excluding those receiving hormone treatment and those with extreme concentration for total T (>4nmol/L for female and 40nmol/L for male) and SHBG (>200nmol/L for female and 150nmol/L for male) (Figure 1). These thresholds were determined by examining the distribution of each sex hormone using quantile-quantile plots in males and females separately (Figure S1).

**Figure 1.**
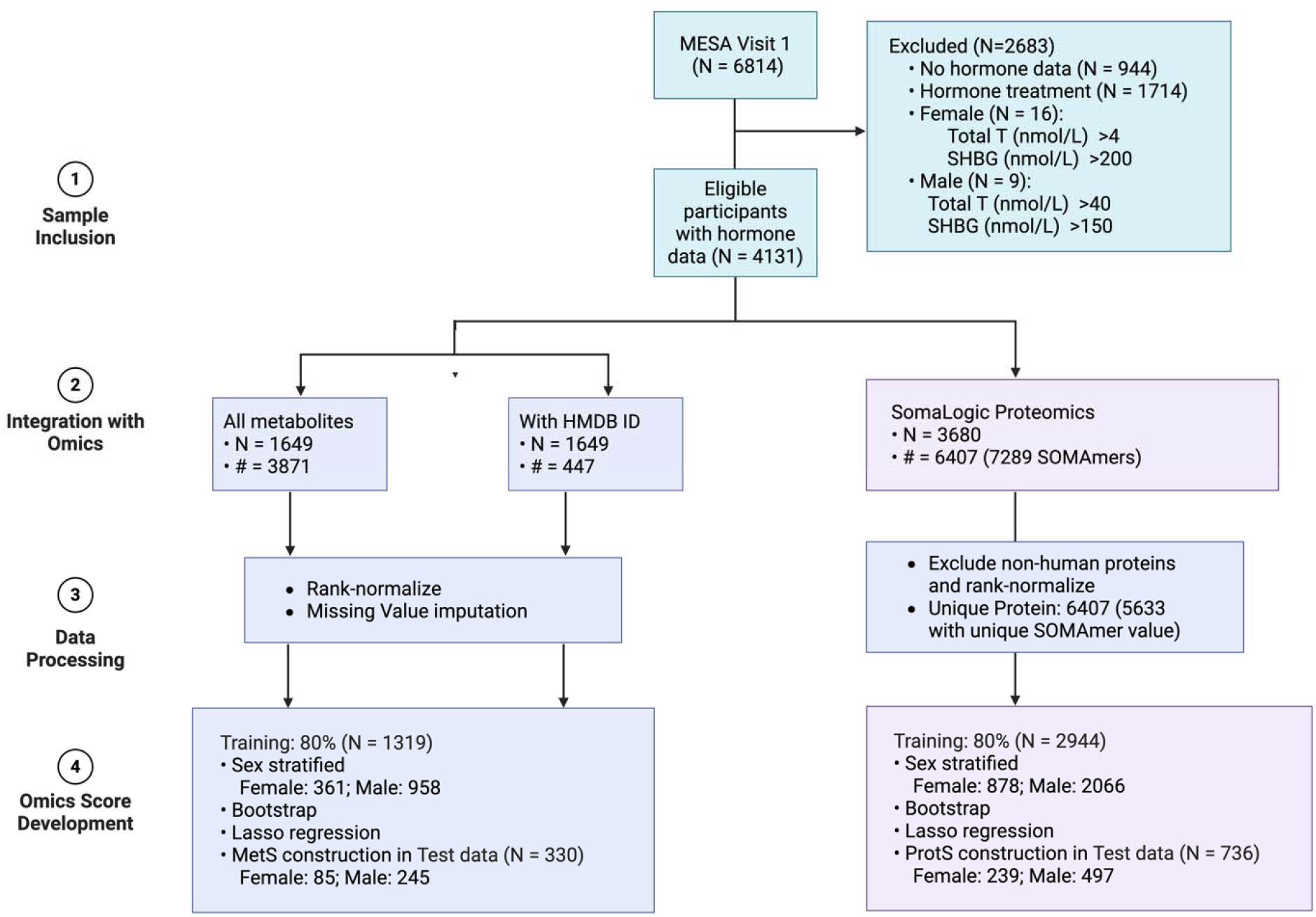
Flowchart for the development of omics-derived hormone scores in MESA, including sample inclusion criteria, omics data integration, preprocessing, and sex-stratified score construction via bootstrap Lasso regression; N: sample size; #: number of omics features.

### SOMAScan™ Proteomic profiling

Plasma protein abundances of 6407 proteins were profiled using 7289 aptamers (SOMAmer) by the SOMAscan platform (SomaLogic, Boulder, CO) following manufacturer’s protocol, as described in detail previously^26^. Standard SOMALogic quality control methods were applied to remove systemic biases, including hybridization control, normalization at SOMAmer, sample, and plate level, and calibration across different assay runs^26,27^. Only human proteins passing quality control were included in this project, with each SOMAmer value rank-normalized to a standard normal distribution with approximate mean 0 and standard deviation 1. SHBG was also captured in the proteomic assay. In total, 3680 consenting MESA participants from baseline had both protein and hormone data, and 3390 and 1394 participants had protein data measured at exam 4 (2005-2007) and 5 (2010-2011), respectively.

### Metabolite profiling

Metabolite profiles were also generated from fasting plasma samples collected at baseline, as well as exams 5 and 6, using liquid chromatography tandem mass spectrometry (LC-MS) methods. Polar metabolites and lipids were profiled using hydrophilic interaction liquid chromatography (HILIC) positive and amide negative modes, and reversed phase C8 chromatography, respectively, as previously describe^22,28,29^. Water-soluble metabolites and lipids were analyzed using Nexera X2 U-HPLC (Shimadzu Corp.; Marlborough, MA) units coupled to a Q Exactive mass spectrometer (Thermo Fisher Scientific; Waltham, MA)^29^. Samples were then normalized across batches and checked for coefficient of variation for quality control^22,29^. Measured peaks, both targeted and untargeted, were processed using HILIC chromatography coupled to a Thermo ID-X Mass Spectrometer (Thermo Fisher Scientific, Waltham, MA) to create a comprehensive tandem MS (MS/MS) library for metabolite identification, with detailed method previously published^22^.

As a result, 3871 metabolites were identified for 3631 MESA participants from baseline, among which 447 metabolites were annotated in Human Metabolome Database (HMDB). For this study we included 1649 participants with available hormone data at baseline. Similar to proteomics data, each metabolite was rank-normalized to have mean 0 and standard deviation 1, and underwent missing value imputation by half of the minimum detected value. After comparing the performance of metabolite scores (MetS) constructed using all available metabolites with those based on HMDB-annotated metabolites, we used the latter in this study because they explained a greater proportion of variance (Table S1).

### The MESA sleep ancillary study

Sleep phenotypes were collected from 2032 participants from Exam 5 (2010-2013) using at-home polysomnography, 7-day wrist actigraphy, and sleep questionnaires, including the Women’s Health Insomnia Rating Scale (WHIIRS) for insomnia and the Epworth Sleepiness Scale (ESS) for daytime sleepiness^30^. In this analysis, the apnea-hypopnea index (AHI) was computed as the average number of apneas and hypopneas with a 3% desaturation per hour of sleep^30^. Mild and moderate to severe obstructive sleep apnea (OSA) was determined using AHI thresholds of 5 and 15, respectively. WHIIRS and ESS were also collected from all Exam 4 participants. This study included 3390 and 1399 participants with both sleep and proteomics data available from Exam 4 and 5, respectively.

### Statistical analyses

We developed protein scores (ProtS) and MetS for each sex hormone, as well as SHBG, in males and females separately, followed by association analysis with corresponding hormones and health phenotypes, including sleep and metabolic disorders. All analyses were conducted using R statistical software^31^.

### Sex stratified protein and metabolite score development

We randomly assigned 80% of the participants from baseline to the training dataset and the remaining to test data (Figure 1). Because hormone distributions deviate from normality, which compromises standard statistical assumptions, we employed 1,000 bootstrap resampling iterations to obtain more robust associations between each hormone and each protein/metabolite, adjusting for age, BMI, race/ethnicity and site in linear regression models. P-values were calculated based on extracted mean and standard deviation of the regression coefficients, and corrected for multiple testing (false discovery rate (FDR) <0.05) to exclude non-associated features. We then applied penalized Lasso regression on analytes (separately for metabolites and proteins) with FDR<0.05 to select key features and eliminate correlated ones, adjusting for the same covariates. Protein and metabolite scores were constructed as weighted sums of Lasso-selected protein/metabolite values.

### Sex stratified assessment of protein and metabolite scores

First, we validated the association between omics scores constructed using the test dataset and their corresponding hormones in males and females separately. In addition to correlation analysis, we estimated the variance explained by each omics score through regression analysis, adjusting for age, BMI, race/ethnicity and study site. Each omics score was plotted against its corresponding hormone in a scatterplot. Functional analysis was conducted using MetaboAnalyst 6.0^32^ with the tight integration method from the Joint Pathway Analysis module to identify functional pathways based on selected metabolites and proteins.

As SHBG is measured in the SOMAScan proteomic assay, we also constructed ProtS excluding SHBG and compared their associations with blood-measured sex hormone levels to those of the original scores.

### Sex stratified association analysis between omics scores and sleep and metabolic phenotypes

All association analyses were performed using simple linear regression with omics scores modeled as exposures, and the phenotype as the outcome, adjusting for the same covariates aforementioned. Metabolic phenotypes including diabetes and hypertension status reported at baseline were also tested for their associations with each omics score, respectively. The omics scores were scaled for association analysis. We further computed ProtS/MetS using data from Exams 4 and 5 and estimated their associations with sleep phenotypes, including log-transformed AHI, mild or moderate-to-severe OSA (Exam 5 only), WHIIRS, and ESS (Exams 4 and 5).

In addition, we fitted Cox proportional hazards models for incident diabetes and hypertension while leveraging repeated measures, i.e. using proteomics measured in exams 1, 4, and 5, and metabolomics measured in exams 1 and 5, at the same model. For this, we used repeated measures from the same individual by incorporating a start-stop interval structure in which the score, age and BMI were updated to the most recent available measurement at the start of each interval (Figure S1) using the R *survival* package v3.5.5^33^. Within-person correlation from multiple intervals was accounted for using robust sandwich variance estimators^33^. When modeling raw hormone measures, available only at baseline, interval-censored proportional hazards models were fitted as sensitivity analysis using the *icenReg* R package v2.0.16^34^ with 200 bootstrap iterations, to model the event as occurring within the interval between the last disease-free visit and the first disease-positive visit. All models were adjusted for age, BMI, and self-reported race, and the Benjamini-Hochberg procedure was applied to control the false discovery rate across hormone scores within each sex-outcome stratum.

## Results

### Demographics characteristics of MESA participants with hormone data

After filtering for hormone therapy and extreme Total T and SHBG values (Figure S2), 4131 (N_Female_= 1289; N_male_=2842) out of 5871 (N_Female_= 2842; N_male_= 3029) participants with available hormone data were included in the study, whose demographic characteristics are summarized in Table 1. Age, BMI, and race/ethnicity composition are comparable between male and female groups. The majority of the female participants (95.8%) had undergone menopause (Table 1). As shown in the quantile-quantile plot (Figure S2), male and female participants showed substantial difference in Total T and Bio T levels.

**Table 1.** Demographic characteristics of all selected MESA participants with hormone data at the baseline exam, stratified by sex.

|  | Overall (n= 4131) | Female (n= 1289) | Male (n= 2842) |
| --- | --- | --- | --- |
| **Age** (mean (SD)) | 63.04 (10.07) | 65.83 (9.35) | 61.78 (10.14) |
| **BMI** (mean (SD)) | 28.20 (5.07) | 29.00 (6.15) | 27.84 (4.45) |

| Race (%) |  |  |  |
| --- | --- | --- | --- |
| **Black** | 1039 (25.2) | 352 (27.3) | 687 (24.2) |
| **Chinese** | 589 (14.3) | 213 (16.5) | 376 (13.2) |
| **Hispanic/Latino** | 1039 (25.2) | 368 (28.5) | 671 (23.6) |
| **White** | 1464 (35.4) | 356 (27.6) | 1108 (39.0) |
| **Gone through menopause (%)** |  |  |  |
| **No** | / | 26 (2.0) | / |
| **YES** | / | 1234 (95.8) | / |
| **Don't know** | / | 28 (2.2) | / |
| **Total T** (mean (SD)) (nmol/L) | 10.61 (7.69) | 1.08 (0.61) | 14.94 (5.09) |
| **Bio T** (mean (SD)) (nmol/L) | 3.86 (2.82) | 0.31 (0.21) | 5.47 (1.80) |
| **Free T** (mean (SD)) (%) | 1.86 (0.55) | 1.53 (0.51) | 2.01 (0.50) |
| **E2** (mean (SD)) (nmol/L) | 0.10 (0.07) | 0.07 (0.09) | 0.12 (0.05) |
| **SHBG** (mean (SD)) (nmol/L) | 47.57 (22.68) | 55.59 (28.34) | 43.94 (18.48) |
| **Diabetes** (%) | 567 (13.7) | 188 (14.6) | 379 (13.3) |
| **Hypertension** (%) | 1837 (44.5) | 667 (51.7) | 1170 (41.2) |

Total T: total testosterone; Bio T: bioavailable testosterone; Free T: free testosterone; E2: estradiol; SHBG: sex hormone binding protein.

### Sex stratified protein and metabolite score development

The numbers of proteins and metabolites selected by the bootstrap filtering and LASSO regression are reported in Table 2. Notably, only 4 proteins and 3 metabolites were associated with Total T in female participants. Still, all constructed ProtS and MetS were significantly associated with their corresponding hormones. The ProtS demonstrated higher variance explained and correlation with the corresponding sex hormones, especially for SHBG (r = 0.81 in females, r = 0.88 in males) and Free T (r = 0.88 in females, r = 0.93 in males), compared to MetS of SHBG (r = 0.60 in females, r = 0.54 in males) and Free T (r = 0.66 in females, r = 0.55 in males) (Table 2, Figure S3). Overall, the ProtS and MetS mirror the internal correlation structures observed among the hormone measures, supporting their use in downstream association modeling (Figure S3). In particular, SHBG and Free T showed a consistently strong negative correlations across both hormone levels and their omics-derived proxies, whereas correlation between Total T and Bio T was substantially higher than that observed between ProtS and MetS. (Figure S3). Excluding SHBG from the ProtS greatly reduced the correlations and variance explained between the testosterone and SHBG ProtS and their corresponding blood-measured levels (Table S2). In contrast, this effect was not observed for E2 in either sex (Table 2; Table S2).

**TABLE 2.** Number of features selected to construct protein and metabolite scores of sex hormones and associations in the test dataset.

|  | 1<sup>st</sup> filtering | Lasso | P-value | Var Explained % |
| --- | --- | --- | --- | --- |
| **FEMALE - PROTEINS (N= 878, #= 7289)** |  |  |  |  |
| **TOTAL T** | 4 | 4 | 4.59e-03 | 4.5 |
| **BIO T** | 2811 | 64 | 2.45e-10 | 21.5 |
| **FREE T** | 4819 | 49 | 1.25e-65 | 77.4 |
| **E2** | 189 | 61 | 1.12e-09 | 12.8 |
| **SHBG** | 4623 | 42 | 3.72e-45 | 65.1 |
| **MALE - PROTEINS (N= 2066, #= 7289)** |  |  |  |  |
| **TOTAL T** | 209 | 89 | 3.50e-57 | 41.6 |
| **BIO T** | 136 | 73 | 3.87e-13 | 19.4 |
| **FREE T** | 1808 | 175 | 1.28e-186 | 85.9 |
| **E2** | 1591 | 113 | 2.05e-14 | 10.4 |
| **SHBG** | 1546 | 80 | 6.64e-137 | 76.8 |
| **FEMALE – METABOLITES (N= 446, #= 447)** |  |  |  |  |
| **TOTAL T** | 3 | 3 | 3.88e-02 | 3.0 |
| **BIO T** | 95 | 24 | 2.54e-03 | 12.8 |
| **FREE T** | 158 | 41 | 4.52e-09 | 45.2 |
| **E2** | 18 | 10 | 2.52e-03 | 15.2 |
| **SHBG** | 155 | 30 | 6.88e-08 | 40.1 |
| **MALE – METABOLITES (N= 1203, #= 447)** |  |  |  |  |
| **TOTAL T** | 134 | 54 | 5.30e-03 | 5.0 |
| **BIO T** | 3 | 3 | 8.93e-03 | 5.6 |
| **FREE T** | 203 | 77 | 4.38e-14 | 28.8 |
| **E2** | 134 | 41 | 6.08e-09 | 13.1 |
| **SHBG** | 208 | 75 | 6.55e-17 | 31.6 |

Total T: total testosterone; Bio T: bioavailable testosterone; Free T: free testosterone; E2: estradiol; SHBG: sex hormone binding protein. Var: variance explained in association analysis in the test data. N: sample size. #: number of analytes included. Considered metabolites are those with known identity and an HMDB identifier (human metabolome database).

The top 5 metabolites and proteins selected for omics scores are reported in Table S3 and S4, with several features being selected for multiple hormones. For instance, dehydroepiandrosterone sulfate was selected across both testosterone and E2 in both sexes, same for androsterone glucuronide across Total T and Bio T (Table S3). However, features selected for Free T and SHBG were more distinct in females compared with males, with greater overlap between Free T and SHBG in males (Table S3-S4). The protein profiles for sex hormones between male and female were largely sex-specific, with SHBG (measured on the proteomics platform) being the only protein selected for both testosterone and SHBG (measured via immunoassay) in both sexes (Table S4).

### Non-linear relationship for SHBG

Upon visualizing omics scores against hormones (Figure S4-S5), omic scores for SHBG exhibited a non-linear relationship with SHBG levels. The inverted U-shaped relationship was confirmed by cubic spline models, where in both males and females, ProtS-SHBG increased with SHBG up to around 2 to 3 units, then slightly declined (Figure S6A). This relationship was less pronounced for MetS-SHBG, which aligned more closely with a linear and positive association (Figure S6B). Moreover, the wider confidence intervals suggest less stable estimate at higher SHBG level for MetS-SHBG (Figure S6B).

### Sex stratified association analysis

#### Omics scores associations with metabolic disorders

Our results indicate that sex hormones affect metabolic disorder risk differently and specifically in males and females. Omic scores for SHBG and Free T were significantly associated with diabetes risk in both males and females, where high ProtS-SHBG (odds ratio (OR_Female_)= 0.4; OR_Male_= 0.67) and lower ProtS-Free T (OR_Female_= 2.5; OR_Male_= 1.49) were protective against diabetes (Figure 2A). Higher ProtS-Bio T (OR= 2.13) increased diabetes risk in females (Figure 2A). In males, both higher ProtS and MetS for SHBG and Total T concentrations were associated with reduced diabetes and hypertension risk, while MetS-Free T increased diabetes risk by approximately 2.5-fold (Figure 2A-2B). On the other hand, hypertension showed no strong directly measured hormone links in women, but higher MetS-E2 (OR= 1.58) and ProtS-Free T (OR= 1.31) are associated with increased hypertension risk in men (Figure 2A-2B). Omics proxy associations mirrored sex hormone associations, except for hypertension where hormone-hypertension associations were non-significant (Figure 2).

**Figure 2.**
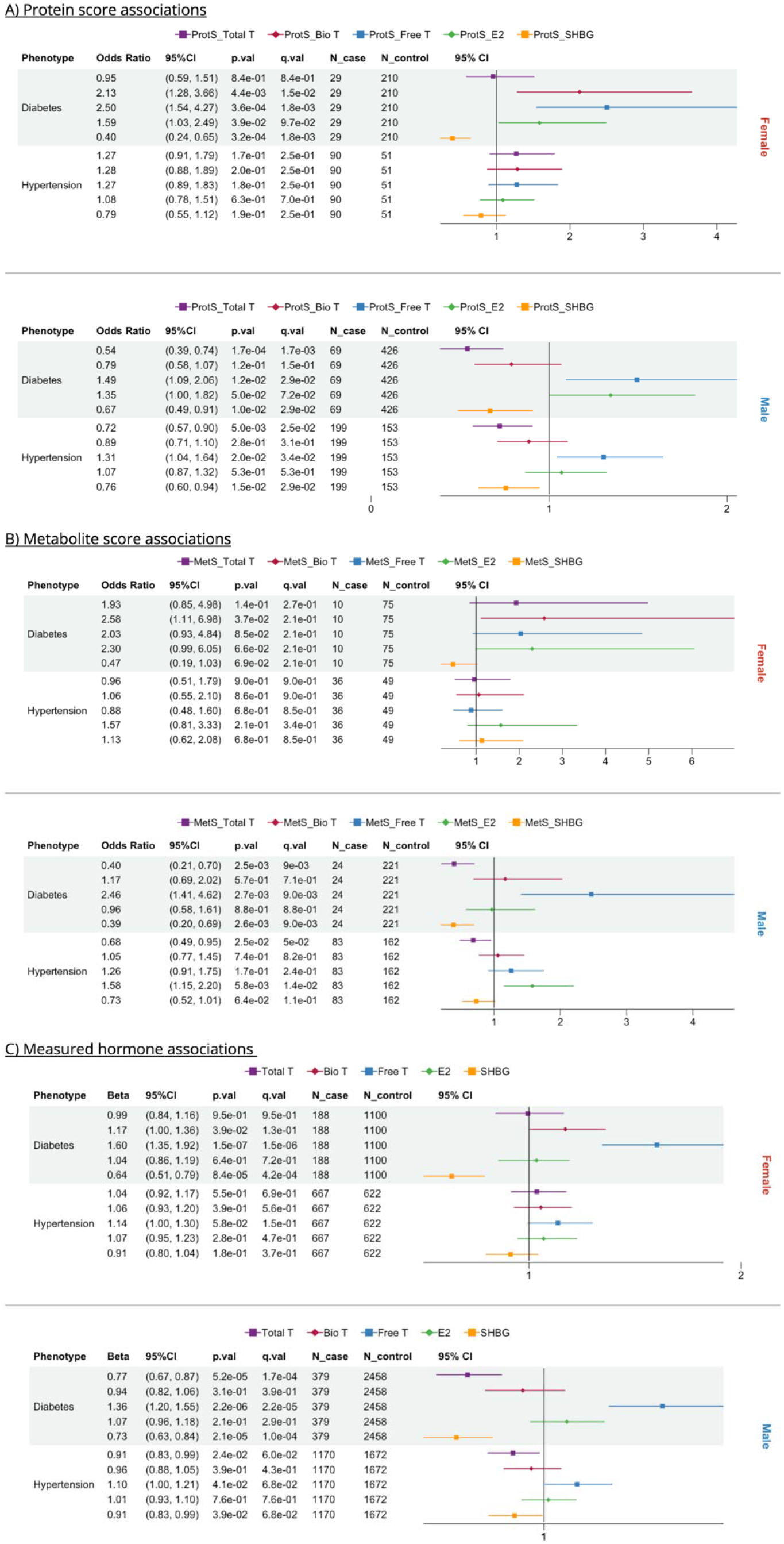
Forest plots of protein (ProtS), metabolite scores (MetS) and sex hormones-based cross-sectional associations with diabetes and hypertension, stratified by A) ProtS; B) MetS; and C) Hormone. Total T: total testosterone; Bio T: bioavailable testosterone; Free T: free testosterone; E2: estradiol; SHBG: sex hormone binding protein.

MetS of Free T (hazard ratio (HR)= 2.59) and SHBG (HR= 0.37) showed strong associations with incident diabetes in female participants, consistent with findings at baseline (Figure 3A, Table S5). No statistically significant associations were observed for MetS in men after FDR correction, despite larger sample size (Figure 3A, Table S5). In contrast, all hormone ProtS were significantly associated with incident diabetes in both sexes, except Total T (Figure 3B). Interestingly, among male participants, ProtS-E2 was the only score showing a stronger association for incident diabetes than in women (HR_Female_= 1.27, HR_Male_= 1.51), and was notably one of only two signals significantly associated with incident hypertension (HR= 1.2), along with Bio T (HR= 1.36) (Figure 3B). While ProtS-Free T (HR= 1.14, p=0.023) and ProtS-SHBG (HR= 0.88, p=0.025) were nominally associated with hypertension in women, neither remained significant after FDR correction (Table S5).

**Figure 3.**
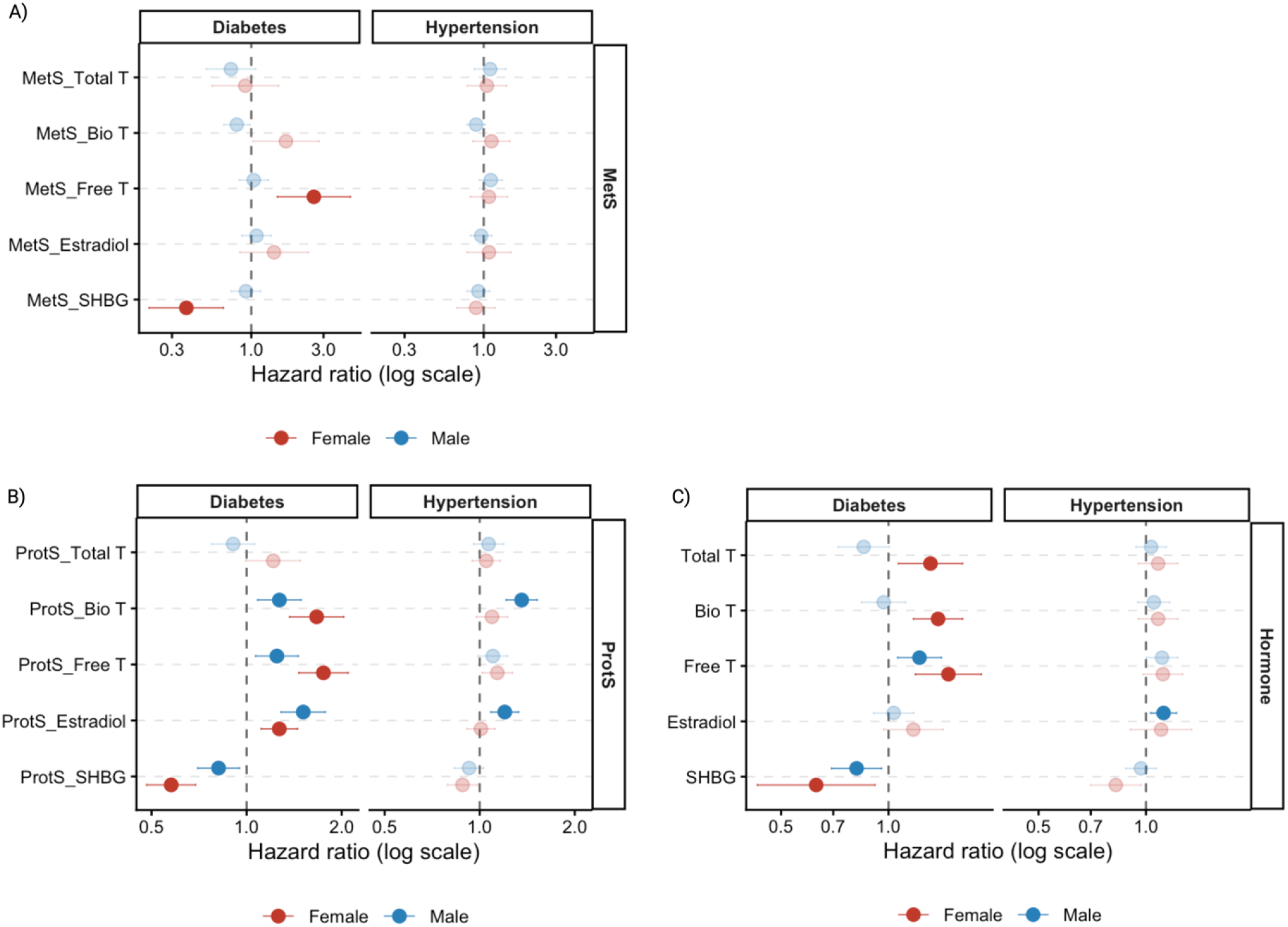
Plots visualizing the associations of metabolite (MetS), protein (ProtS) scores, and measured sex hormones with incident diabetes and hypertension, by A) MetS; B) ProtS and C) sex hormones. Total T: total testosterone; Bio T: bioavailable testosterone; Free T: free testosterone; E2: estradiol; SHBG: sex hormone binding protein.

Results from interval-censored model using measured hormones corroborated the ProtS and MetS findings (Figure 3C, Table S6). In particular, Total T exhibited sex-specific associations with incident diabetes (HR_Female_= 1.31, HR_Male_= 0.85), with a borderline association observed in men (p= 0.057) (Table S6). E2 remained significantly associated with incident hypertension in men only (HR= 1.12) (Figure 3C, Table S6). Furthermore, adding MetS and ProtS improved *discrimination compared* to the base model including only clinical covariates (age, BMI and race), with MetS increasing the C-statistic by up to 0.08 in female participants (Table S5).

#### Omics scores associations with sleep phenotypes

Figure 4 shows the sleep phenotypes with statistically significant associations with sex hormone–related omics scores at Exam 5. Table S7 and Table S8 report the coefficients and p-values for all tested sleep phenotypes. For female participants, while ProtS of sex hormones showed no associations with sleepiness or insomnia symptoms (ESS, WHIIRS, or insomnia), those of Free T and Bio T were positively associated with more severe OSA as measured by a higher AHI and OSA case status (Figure 4B, Table S8). Similar to metabolic disorders, omics scores for SHBG showed a moderate to strong protective effect for AHI in women (β= -0.17, Table S8) and OSA (AHI >5) in both sexes (Figure 4). In men, higher MetS-Total T was also associated with decreased risk for OSA (AHI >5), and higher MetS-E2 (OR= 1.32) was linked to increased insomnia risk (Figure 4A). Interestingly, only ProtS in females and MetS in males showed significant associations with sleep phenotypes (Figure 4). MESA participants at Exam 4 only had only questionnaire-based sleep phenotypes collected (WHIIRS and ESS) and measured proteomics, and neither showed significant associations with ProtS of the 5 sex hormones despite the increased sample size (Figure S7).

**Figure 4.**
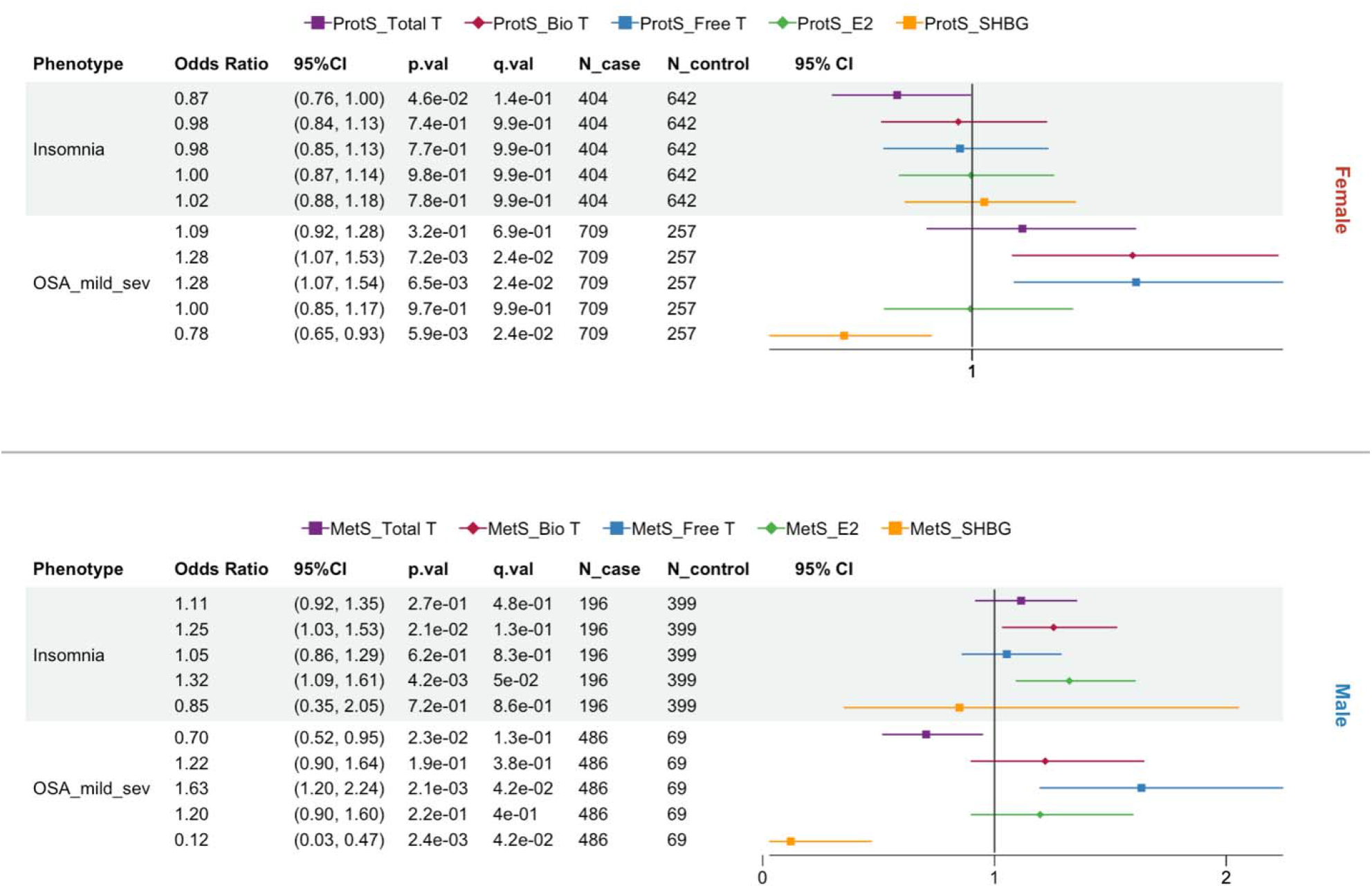
Forest plots of protein (ProtS) and metabolite scores (MetS) based statistically significant associations across sleep phenotypes, stratified by Female and Male participants. Total T: total testosterone; Bio T: bioavailable testosterone; Free T: free testosterone; E2: estradiol; SHBG: sex hormone binding protein. WHIIRS: Women’s Health Insomnia Rating Scale; OSA: obstructive sleep apnea.

### Pathway analysis

MetaboAnalyst identified 8 and 1 biological pathways (FDR<0.05) for Free T and Bio T, respectively and none for SHBG, in females, while queries based on features selected for other hormones revealed no statistically significant pathways (Table 3). In males, 2 and 8 functional pathways were identified for Total and Free T, respectively (Table 3). No significant functional pathways were identified for Bio T in men, whereas features associated with SHBG had 14 pathways identified, which exceeded that for the sex hormones (Table 3).

**Table 3.** Functional pathways identified combining selected proteins and metabolites using Metaboanalyst.

| PATHWAY NAME | # FEATURE | FDR | HORMONE |
| --- | --- | --- | --- |
| **FEMALE** |  |  |  |
| Central carbon metabolism in cancer | 5, 7 | 2.6E-02, 1.3E-04 | Bio T, Free T |
| Protein digestion and absorption | 8 | 1.3E-04 | Free T |
| Mineral absorption | 6 | 4.9E-04 | Free T |
| Aminoacyl-trna biosynthesis | 6 | 1.8E-03 | Free T |
| ABC transporters | 7 | 1.8E-03 | Free T |
| Complement and coagulation cascades | 5 | 1.5E-02 | Free T |
| Vitamin digestion and absorption | 4 | 1.6E-02 | Free T |
| D-amino acid metabolism | 4 | 2.0E-02 | Free T |
| **MALE** |  |  |  |
| Valine, leucine and isoleucine biosynthesis | 3, 3 | 2.6E-03, 2.0E-02 | Total T, SHBG |
| Pantothenate and CoA biosynthesis | 3, 4 | 2.4E-02, 1.6E-02 | Total T, SHBG |
| Central carbon metabolism in cancer | 10, 7, 9 | 1.2E-04, 5.9E-03, 1.2E-05 | Free T, E2, SHBG |
| D-Amino acid metabolism | 8, 7 | 3.3E-04, 1.E-04 | Free T, SHBG |
| Complement and coagulation cascades | 8 | 5.5E-03 | Free T |
| Glycine, serine and threonine metabolism | 7, 6 | 5.5E-03, 2.E-03 | Free T, SHBG |
| ABC transporters | 9, 8 | 1.6E-02, 2.0E-03 | Free T, SHBG |
| Protein digestion and absorption | 8 | 1.8E-02 | Free T |
| Glyoxylate and dicarboxylate metabolism | 6, 5 | 3.5E-02, 1.6E-02 | Free T, SHBG |
| Alanine, aspartate and glutamate metabolism | 5, 5 | 3.6E-02, 4.0E-03 | Free T, SHBG |
| Protein digestion and absorption | 8 | 8.2E-04 | SHBG |
| Aminoacyl-tRNA biosynthesis | 6 | 6.5E-03 | SHBG |
| Fat digestion and absorption | 4 | 1.8E-02 | SHBG |
| Linoleic acid metabolism | 4 | 2.0E-02 | SHBG |
| Cholesterol metabolism | 4 | 2.0E-02 | SHBG |
| Phenylalanine metabolism | 4 | 2.3E-02 | SHBG |

The table provides, for each sex groups, pathway identified by selected proteins and metabolites by LASSO regressions for each sex hormone. When a pathway was highlighted for more than one hormone, the information is displayed in a single row using commas as separators of the information corresponding to each of the hormones. # of features is the number of analytes (metabolites or proteins) representing the pathway among the selected metabolites and proteins, FDR is the false-discovery rate of the enrichment hypothesis test.
Total T: total testosterone; Bio T: bioavailable testosterone; Free T: free testosterone; E2: estradiol; SHBG: sex hormone binding protein.

## Discussion

We developed sex-specific proteomics and metabolomics scores for sex hormones in MESA using a feature selection approach. The developed omics scores showed strong associations with their corresponding hormones, and revealed sex-specific associations with sleep phenotypes and incident metabolic disorders. Unlike the sex hormones, SHBG exhibited a non-linear relationship with its omics score, particularly with ProtS. In general, ProtS explained a higher percentage of variance in sex hormones and SHBG than MetS, and exhibited stronger prospective associations with incident cardiometabolic diseases, especially in women. One potential reason for more significant associations observed for sex hormone omic-scores in women than in men was the greater interindividual variability in endogenous sex hormone levels in post-menopausal women^35,36^, consistent with the heightened cardiometabolic vulnerability following estrogen withdrawal^37^.

Hormone levels and omic proxies of Free T, estradiol, and SHBG were highly consistent in their associations with metabolic disorders (Figure 2-3). ^3,38^The observed protective effect of SHBG against metabolic disorders aligns with previous studies^5,39^. Nevertheless, it is also possible that SHBG levels are influenced by high insulin level and hypertension^40–42^. In contrast to previous research where E2 levels were found to be positively correlated with pulmonary arterial hypertension in postmenopausal women and men, along with lower levels of dehydroepiandrosterone-sulfate (DHEA-S)^38^, our developed omics scores of Bio T and E2 were associated with increased incident hypertension risk only in men. In addition, such associations were not observed for these hormones at baseline, when the cohort was younger and hypertension was less common, and perhaps due to lower power resulted from a single time-point measurement (whereas omics were measured up to three times). Accordingly, our results suggest a comparable link between E2 and systemic hypertension, manifesting strong sex-specific associations and sensitivity to hypoxia^43,44^. Although DHEA-S is selected by Lasso for MetS-E2 in females only, its specific relationship with E2 and these hypertensive phenotypes between the two sexes warrants further functional verification.

Most commonly used free T calculations method are strongly correlated with free T measured by reference methods^45^. However, the theoretical basis underlying most commonly used free T calculation methods is now known to be flawed—particularly in women^45^. This is because free T calculation relies on SHBG concentrations^24^, and E2 can alter the binding of testosterone to SHBG. In women, circulating E2 levels are up to tenfold higher and substantially more variable than those of men, where the impact of E2 on free T is lower. Accordingly, the calculated Free T results should be interpreted cautiously, and the possibility that they actually reflect the impact of SHBG should be considered, particularly as majority of the pathways pertinent to Free T were also identified for SHBG in males (Table 3). In contrast to previous research^3^, both blood level and omics scores of Free T showed positive associations with higher risk of diabetes in both sexes. Considering the challenges in accurately quantifying Free T, additional validation is needed to confirm its relationship with health outcomes in both sexes.

Recent evidence highlights that SHBG may have effects that are not related to simply being a carrier protein^39^, which also aligns with the observed associations with metabolic and sleep outcomes in the current study. Consistent with a meta-analysis of 24 studies in women with polycystic ovary syndrome ^46^, ProtS-SHBG had a protective association with AHI severity level and mild to severe OSA in women. Low SHBG levels would result in higher Free and Bio T levels, whose relationship with OSA in females is also reflected by ProtS (Figure 4). In men, MetS of SHBG and Free T exhibited the same direction of association with mild to severe OSA risk as in women, but no significant association was observed for Total T (Figure 4). That said, the sex-dependent prevalence in OSA associated with age-related reduction in Free T may be closely related to SHBG. In both sexes, reduced SHBG concentrations may be associated with increased OSA risk via insulin resistance, with potential effects on ventilatory control and pharyngeal muscle function ^47,48^.

Our results showed significant associations between OSA and omics proxies for both SHBG and Free T in opposite directions across men and women, but not for Total T, reiterating the important role played by SHBG in OSA pathology. Nevertheless, the exact relationships and mechanisms underlying OSA subtype heterogeneity and hormonal changes at different life stages require further verification in large population studies. Furthermore, higher omics scores of E2 were found associated with increased insomnia risk in males only. However, previous studies have shown conflicting results regarding the effect of E2 on insomnia^49,50^, which warrants further study to establish its long-term and short-term impact on insomnia in men.

The top features selected for SHBG manifested sex-specific profiles, sharing only 7 proteins and 0 metabolites between the two sexes (Supplementary data). These sex-specific proteins and metabolites may serve as potential biomarkers behind testosterone vs estrogen dominant physiology modulated by SHBG. For instance, insulin like growth factor binding protein 1 (IGFBP1) and mucosa associated lymphoid tissue lymphoma translocation protein 1 (MALT1) suggested tight connections of SHBG with insulin resistance^51^ and immune suppression^52,53^ in women, possibly through modulation of Free T and E2 content, which is influenced by thyroxine by regulating SHBG production^54^. In males, selected systemic inflammation and OSA severity markers, matrix metalloproteinase (MMP2 and MMP9), highlight a potential link between SHBG and hypoxemia-induced oxidative stress^55,56^. With the increasing availability of omics data in large cohorts and rapid advances in analytical approaches, a key emerging direction is to pinpoint key biomarkers that drive sex differences in multifaceted sleep disorders through integrated multi-omics frameworks^57^.

To the best of our knowledge, our study is the first to develop omics scores for sex hormones and SHBG and investigate their associations with metabolic and sleep phenotypes in men and women with a mean age between 60 and 65 years. Our developed omics scores, along with their associations with sleep outcomes, could be limited to this particular age group, given that sex hormone levels undergo substantial changes across different stages of life. Another limitation is that exact diagnosis dates for incident diabetes and hypertension were unavailable, and assigning study visit date as event timing in the Cox models could attenuate estimated hazard ratios toward the null. To mitigate this limitation, interval-censored proportional hazards models were fitted for measured hormones at baseline. The consistency of findings across both analytical approaches supports the robustness of the observed associations. Accordingly, it will be of great interest to validate the omics scores in populations of different age groups, including individuals of reproductive age, and to evaluate their potential as predictive risk factor for diseases manifesting sex-specific prevalence, particularly the role of E2 in men. This is important as generalization of omics scores is often limited by different assay versions and analytical platforms^58^. Given that both OSA and insomnia have been associated with metabolic dysfunction^59^, our results warrant future research to investigate the hormonal mechanisms that mediate bidirectional relationships between metabolic diseases and sleep disorders.

## Conclusions

Our findings demonstrate that these omics scores serve as reliable proxies for sex hormone levels, which could serve to represent hormonal status in large-scale cohort studies where direct measurement may be impractical. The convergence of Bio and Free T as risk factors and SHBG as protective marker across both baseline and follow-up measurements strengthens the biological plausibility of androgen-mediated cardiometabolic risk. The sex-specific molecular profiles and functional pathways identified provide insights into the biological mechanisms, and indicate that the hormone omic scores reflect stable biological pathways relevant to cardiometabolic risk, rather than transient fluctuations.

## Data Availability

MESA phenotype data are available from dbGaP according to the MESA accession: MESA: phs000209.

## Acknowledgements

The MESA projects are conducted and supported by the National Heart, Lung, and Blood Institute (NHLBI) in collaboration with MESA investigators. Support for MESA is provided by contracts 75N92025D00022, 75N92020D00001, HHSN268201500003I, N01-HC-95159, 75N92025D00026, 75N92020D00005, N01-HC-95160, 75N92020D00002, N01-HC-95161, 75N92025D00024, 75N92020D00003, N01-HC-95162, 75N92025D00027, 75N92020D00006, N01-HC-95163, 75N92025D00025, 75N92020D00004, N01-HC-95164, 75N92025D00028, 75N92020D00007, N01-HC-95165, N01-HC-95166, N01-HC-95167, N01-HC-95168, N01-HC-95169, UL1-TR-000040, UL1-TR-001079, UL1-TR-001420, UL1TR001881, U01DK140761, and R01HL105756. MESA proteomic data were generated under NIH 1R01HL159081. The MESA Sleep Ancillary studies were funded by NIH-NHLBI R01HL098433. The authors thank the MESA participants and the MESA investigators and staff for their valuable contributions. A full list of participating MESA investigators and institutions can be found at http://www.mesa-nhlbi.org. Molecular data for the Trans-Omics in Precision Medicine (TOPMed) program was supported by the National Heart, Lung and Blood Institute (NHLBI). Metabolomics for “NHLBI TOPMed: MESA and MESA Family AA-CAC” (phs001416.v4.p1) was performed at the Broad Institute and Beth Israel Metabolomics Platform (HHSN268201600038I). Core support including centralized genomic read mapping and genotype calling, along with variant quality metrics and filtering were provided by the TOPMed Informatics Research Center (3R01HL-117626-02S1; contract HHSN268201800002I). Core support including phenotype harmonization, data management, sample-identity QC, and general program coordination were provided by the TOPMed Data Coordinating Center (R01HL-120393; U01HL-120393; contract HHSN268201800001I). We gratefully acknowledge the studies and participants who provided biological samples and data for TOPMed.

## Author contribution

Z.W conduct analysis, write and revise the manuscript; Y.Zhang process metabolomic data, review and revise the manuscript; A.Wood, M.Benson, K.Taylor, X.Guo, S.Rich, R.Gerszten, S.Redline, J.Rotter, P.Ganz, R.Deo, R.Dubin prepare and curate the MESA data; review and revise the manuscript; P.Liu provide guidance on analysis, review and revise the manuscript; T.Sofer conceive the study and supervise the analysis; review and revise the manuscript.

## Ethics approval and consent to participate

This work was approved by the Beth Israel Deaconess Medical Center Committee on Clinical Investigations, protocol #2023P000541. All MESA participants provided written informed consent, and the study was approved by the Institutional Review Boards at The Lundquist Institute (formerly Los Angeles BioMedical Research Institute) at Harbor-UCLA Medical Center, University of Washington, Wake Forest School of Medicine, Northwestern University, University of Minnesota, Columbia University, and Johns Hopkins University.

## Data availability

MESA phenotype data are available through an approved Data Access Requests in dbGaP accession phs000209, and metabolomics are similarly available through dbGaP accession phs001416. MESA phenotypes, metabolomics, and proteomics data are also available via a data use agreement from the MESA Coordinating Center (see investigator website https://www.mesa-nhlbi.org/).

