## Supplementary Files for "Sex-specific omics scores of sex hormones and associations with metabolic and sleep disorders" (from another version of this preprint: version 1, 2026-04-28)

### Supplementary Tables

#### Table S1. Number of metabolites selected to construct metabolite scores (MetS) of sex hormones using all metabolites and those annotated in Human Metabolome Database (HMDB).

|  | 1st filtering | Lasso | P-value | Var Explained % |
| --- | --- | --- | --- | --- |
| FEMALE – All Metabolites (N= 446, #= 3871) | | | | |
| Total TST | 6 | 4 | 5.96e-02 | 4.4 |
| Bioavailable TST | 203 | 40 | 1.01e-07 | 36.6 |
| Free TST | 706 | 73 | 5.92e-06 | 28.4 |
| Estradiol | 76 | 36 | 3.36e-05 | 22.4 |
| SHBG | 701 | 75 | 5.52e-06 | 28.3 |
| *Female – HMDB Metabolites (N= 446, #= 447) | | | | |
| Total TST | 3 | 3 | 3.88e-02 | 3.0 |
| Bioavailable TST | 95 | 24 | 2.54e-03 | 12.8 |
| Free TST | 158 | 41 | 4.52e-09 | 45.2 |
| Estradiol | 18 | 10 | 2.52e-03 | 15.2 |
| SHBG | 155 | 30 | 6.88e-08 | 40.1 |
| MALE – All Metabolites (N= 1203, #= 3871) | | | | |
| Total TST | 364 | 94 | 3.30e-01 | 0.7 |
| Bioavailable TST | 11 | 10 | 6.00e-01 | 0.1 |
| Free TST | 934 | 148 | 1.25e-11 | 18.0 |
| Estradiol | 1039 | 168 | 6.01e-09 | 14.9 |
| SHBG | 935 | 161 | 1.89e-12 | 18.9 |
| *MALE – HMDB Metabolites (N= 1203, #= 447) | | | | |
| Total TST | 134 | 54 | 5.30e-03 | 5.0 |
| Bioavailable TST | 3 | 3 | 8.93e-03 | 5.6 |
| Free TST | 203 | 77 | 4.38e-14 | 28.8 |
| Estradiol | 134 | 41 | 6.08e-09 | 13.1 |
| SHBG | 208 | 75 | 6.55e-17 | 31.6 |

* Indicate the MetS selected for this study.

| Table S2. Correlations and Associations of Sex Hormone Protein Scores (ProtS) excluding SHBG With Corresponding Hormone Levels in the Test Dataset | | | |
| --- | --- | --- | --- |
|  | **Correlation with corresponding ProtS** | **P-value** | **Var Explained %** |
| FEMALE - Proteins (N= 878, #= 7289) | | | |
| Total T | 0.21 | 0.0046 | 4.51 |
| Bio T | 0.44 | 4.88E-09 | 19.30 |
| Free T | 0.38 | 2.00E-05 | 14.50 |
| E2 | 0.36 | 1.12E-09 | 12.79 |
| SHBG | 0.34 | 6.45E-05 | 11.52 |
| MALE - Proteins (N= 2066, #= 7289) | | | |
| Total T | 0.46 | 4.08E-18 | 21.47 |
| Bio T | 0.44 | 3.87E-13 | 19.39 |
| Free T | 0.42 | 2.89E-16 | 18.04 |
| E2 | 0.32 | 2.05E-14 | 10.44 |
| SHBG | 0.27 | 1.38E-08 | 7.24 |

Total T: total testosterone; Bio T: bioavailable testosterone; Free T: free testosterone; E2: estradiol; SHBG: sex hormone binding protein. Var: variance explained in association analysis in the test data. N: sample size. #: number of analytes included.

| Table S3. Top 5 metabolites selected for sex hormones by sex | | | |
| --- | --- | --- | --- |
| FEATURE | **Name** | **HORMONE** | |
| MALE | | | |
| HMDB0002829 | Androsterone glucuronide | Total T, Bio T, E2 | |
| HMDB0054096 | Tracylglycerol (54:10) | Total T | |
| HMDB0001867 | 4-Aminohippuric acid | Total T | |
| HMDB0000092 | Dimethylglycine | Total T | |
| HMDB0060564 | Carboxy-ibuprofen | Total T | |
| HMDB0001032 | Dehydroepiandrosterone sulfate | Bio T, Free T, E2 | |
| HMDB0000774 | Pregnenolone sulfate | Bio T | |
| HMDB0012103 | Behenoyl sphingomyelin | Free T, SHBG | |
| HMDB0010169 | C16 Sphingomyelin | Free T, SHBG | |
| HMDB0240613 | Sphingomyelin (D18:1/16:1) | Free T | |
| HMDB0002005 | Methionine sulfoxide | Free T | |
| HMDB0000123 | Glycine | E2 | |
| HMDB0013713 | N-Acetyltryptophan | E2 | |
| HMDB0000248 | Thyroxine | E2 | |
| HMDB0007883 | Phosphatidylcholine (14:0/20:4) | SHBG | |
| HMDB0000982 | 5-Methylcytidine | SHBG | |
| HMDB0001348 | Stearoyl sphingomyelin | SHBG | |
| FEMALE | | | |
| HMDB0011478 | Lysophosphatidylethanolamine (18:3) | | Total T, Bio T |
| HMDB0001032 | Dehydroepiandrosterone sulfate | | Total T, E2 |
| HMDB0002829 | Androsterone glucuronide | | Total T, Bio T |
| HMDB0042098 | Tracylglycerol (43:1) | | Bio T |
| HMDB0008511 | Phosphatidylcholine (40:10) | | Bio T |
| HMDB0240212 | Dimethylguanidino valeric acid | | Bio T, Free T, SHBG |
| HMDB0004952 | N-(Docosanoyl)ceramide | | Free T, SHBG |
| HMDB0010390 | Lysophosphatidylcholine (20:0) | | Free T, SHBG |
| HMDB0000248 | Thyroxine | | Free T, SHBG |
| HMDB0008057 | Phosphatidylcholine (18:0/22:6) | | Free T |
| HMDB0000191 | L-Aspartic acid | | E2 |
| HMDB0005060 | Eicosadienoic acid | | E2 |
| HMDB0008036 | Distearoyl phosphatidylcholine | | E2 |
| HMDB0000884 | Ribothymidine | | E2 |
| HMDB0013122.1 | Lysophosphatidylcholine (18:1) | | SHBG |

Total T: total testosterone; Bio T: bioavailable testosterone; Free T: free testosterone; E2: estradiol; SHBG: sex hormone binding protein.

| Table S4. Top proteins selected for sex hormones by sex | | | |
| --- | --- | --- | --- |
| FEATURE | **Name** | **HORMONE** | |
| MALE | | | |
| P04278 | Sex hormone-binding globulin | Total T, Free T, SHBG | |
| P09529 | Inhibin beta B chain | Total T, Bio T, E2 | |
| Q8IW52 | SLIT and NTRK-like protein 4 | Total T | |
| P41159 | Leptin | Total T, Bio T | |
| Q8WTQ1 | Beta-defensin 104 | Total T, Bio T | |
| P15090 | Fatty acid-binding protein, adipocyte | Bio T | |
| P51460 | Insulin-like 3 | Bio T | |
| Q5TAT6 | Collagen alpha-1(XIII) chain | Free T | |
| P07225 | Vitamin K-dependent protein S | Free T | |
| P0C0P6 | Neuropeptide S | Free T | |
| O14511 | Pro-neuregulin-2 | Free T | |
| O76076 | WNT1-inducible-signaling pathway protein 2 | E2 | |
| P43652 | Afamin | E2 | |
| Q03167 | Transforming growth factor beta receptor type 3 | E2 | |
| O75771 | DNA repair protein RAD51 homolog 4 | E2 | |
| P02774 | Vitamin D-binding protein | SHBG | |
| Q9H2X3 | C-type lectin domain family 4 member M | SHBG | |
| P08697 | Alpha-2-antiplasmin | SHBG | |
| O75882 | Attractin | SHBG | |
| FEMALE | | | |
| P00750 | Tissue-type plasminogen activator | | Total T |
| Q07075 | Glutamyl aminopeptidase | | Total T |
| P22888 | Lutropin-choriogonadotropic hormone receptor | | Total T, Bio T |
| P01266 | Thyroglobulin | | Total T |
| P04278 | Sex hormone-binding globulin | | Bio T, Free T, SHBG |
| P00750 | Tissue-type plasminogen activator | | Bio T |
| P22888 | Lutropin-choriogonadotropic hormone receptor | | Bio T |
| Q13822 | Ectonucleotide pyrophosphatase | | Bio T |
| P61371 | Insulin gene enhancer protein ISL-1 | | Free T, SHBG |
| P08833 | Insulin-like growth factor-binding protein 1 | | Free T |
| Q9UDY8 | Mucosa-associated lymphoid tissue lymphoma  translocation protein 1 | | Free T, SHBG |
| Q9NWS6 | Protein FAM118A | | Free T |
| Q6WN34 | Chordin-like protein 2 | | E2 |
| Q6FHJ7 | Secreted frizzled-related protein 4 | | E2 |
| Q9HCK4 | Roundabout homolog 2 | | E2 |
| P43652 | Afamin | | E2 |
| P53582 | Methionine aminopeptidase 1 | | E2 |
| P05156 | Complement factor I | | SHBG |
| Q13506 | NGFI-A-binding protein 1 | | SHBG |

Total T: total testosterone; Bio T: bioavailable testosterone; Free T: free testosterone; E2: estradiol; SHBG: sex hormone binding protein.

| Table S5. Association analysis for metabolite (MetS) and protein (ProtS) scores of sex hormones and metabolic disorders using counting process cox hazard model | | | | | | |
| --- | --- | --- | --- | --- | --- | --- |
| Phenotype | **Hazard ratio** | **p-value** | **95% CI** | **pval_fdr** | **C_statistic** | **Omics measure** |
| MetS - Female | | | | | | |
| Hypertension | 1.049 | 0.756 | (0.78‚ 1.41) | 0.756 | 0.572 | Mets. Total T |
| Hypertension | 1.124 | 0.415 | (0.85, 1.49) | 0.756 | 0.567 | Mets. Bio T |
| Hypertension | 1.082 | 0.586 | (0.82, 1.44) | 0.756 | 0.562 | Mets. Free T |
| Hypertension | 1.083 | 0.641 | (0.78‚ 1.51) | 0.756 | 0.587 | Mets. E2 |
| Hypertension | 0.89 | 0.428 | (0.67‚ 1.19) | 0.756 | 0.561 | Mets. SHBG |
| Diabetes | 0.913 | 0.725 | (0.55‚ 1.52) | 0.725 | 0.736 | Mets. Total T |
| Diabetes | 1.695 | 0.0404 | (1.02‚ 2.81) | 0.067 | 0.77 | Mets. Bio T |
| Diabetes | 2.59 | 0.00081 | (1.48‚ 4.52) | 0.002 | 0.82 | Mets. Free T |
| Diabetes | 1.415 | 0.193 | (0.84‚ 2.39) | 0.241 | 0.772 | Mets. E2 |
| Diabetes | 0.373 | 0.00061 | (0.21‚ 0.66) | 0.002 | 0.825 | Mets. SHBG |
| MetS - Male | | | | | | |
| Hypertension | 1.102 | 0.43 | (0.87‚ 1.41) | 0.54 | 0.587 | Mets. Total T |
| Hypertension | 0.891 | 0.102 | (0.78‚ 1.02) | 0.51 | 0.586 | Mets. Bio T |
| Hypertension | 1.113 | 0.229 | (0.94‚ 1.33) | 0.54 | 0.579 | Mets. Free T |
| Hypertension | 0.961 | 0.632 | (0.82‚ 1.13) | 0.63 | 0.586 | Mets. E2 |
| Hypertension | 0.921 | 0.357 | (0.77‚ 1.1) | 0.54 | 0.579 | Mets. SHBG |
| Diabetes | 0.734 | 0.108 | (0.5‚ 1.07) | 0.27 | 0.694 | Mets. Total T |
| Diabetes | 0.803 | 0.0311 | (0.66‚ 0.98) | 0.156 | 0.688 | Mets. Bio T |
| Diabetes | 1.039 | 0.735 | (0.83‚ 1.3) | 0.735 | 0.682 | Mets. Free T |
| Diabetes | 1.083 | 0.479 | (0.87‚ 1.35) | 0.599 | 0.687 | Mets. E2 |
| Diabetes | 0.922 | 0.475 | (0.74‚ 1.15) | 0.599 | 0.683 | Mets. SHBG |
| ProtS - Female | | | | | | |
| Hypertension | 1.049 | 0.372 | (0.95, 1.16) | 0.47 | 0.644 | Prots. Total T |
| Hypertension | 1.093 | 0.128 | (0.98, 1.23) | 0.21 | 0.644 | Prots. Bio T |
| Hypertension | 1.137 | 0.023 | (1.02, 1.27) | 0.064 | 0.643 | Prots. Free T |
| Hypertension | 1.009 | 0.859 | (0.91, 1.12) | 0.86 | 0.644 | Prots. E2 |
| Hypertension | 0.882 | 0.0254 | (0.79, 0.99) | 0.064 | 0.643 | Prots. SHBG |
| Diabetes | 1.214 | 0.0582 | (0.99, 1.48) | 0.058 | 0.726 | Prots. Total T |
| Diabetes | 1.666 | 4.17E-07 | (1.37, 2.03) | 6.95E-07 | 0.754 | Prots. Bio T |
| Diabetes | 1.751 | 1.19E-09 | (1.46, 2.1) | 4.38E-09 | 0.753 | Prots. Free T |
| Diabetes | 1.269 | 0.00045 | (1.11, 1.45) | 0.00056 | 0.731 | Prots. E2 |
| Diabetes | 0.577 | 1.75E-09 | (0.48, 0.69) | 4.38E-09 | 0.753 | Prots. SHBG |
| ProtS - Male | | | | | | |
| Hypertension | 1.067 | 0.246 | (0.96, 1.19) | 0.246 | 0.605 | Prots. Total T |
| Hypertension | 1.358 | 1.24E-07 | (1.21, 1.52) | 6.20E-07 | 0.628 | Prots. Bio T |
| Hypertension | 1.1 | 0.0845 | (0.99, 1.23) | 0.14 | 0.606 | Prots. Free T |
| Hypertension | 1.2 | 0.00043 | (1.08, 1.32) | 0.0011 | 0.614 | Prots. E2 |
| Hypertension | 0.925 | 0.156 | (0.83, 1.03) | 0.195 | 0.606 | Prots. SHBG |
| Diabetes | 0.906 | 0.221 | (0.77, 1.06) | 0.221 | 0.666 | Prots. Total T |
| Diabetes | 1.27 | 0.0031 | (1.08, 1.49) | 0.0078 | 0.664 | Prots. Bio T |
| Diabetes | 1.247 | 0.0054 | (1.07, 1.46) | 0.00895 | 0.669 | Prots. Free T |
| Diabetes | 1.511 | 5.74E-07 | (1.29, 1.78) | 2.87E-06 | 0.696 | Prots. E2 |
| Diabetes | 0.815 | 0.0084 | (0.7, 0.95) | 0.011 | 0.669 | Prots. SHBG |

C-statistic using only clinical covariates (Age, BMI, Race): MetS-Female (Hypertension: 0.581, Diabetes: 0.739); MetS-Male (Hypertension: 0.584, Diabetes: 0.682); ProtS-Female (Hypertension: 0.644, Diabetes: 0.716); ProtS-Male (Hypertension: 0.605, Diabetes: 0.663).

Total T: total testosterone; Bio T: bioavailable testosterone; Free T: free testosterone; E2: estradiol; SHBG: sex hormone binding protein.

| Table S6. Association analysis for sex hormones and metabolic disorders using interval-censored model | | | | | |
| --- | --- | --- | --- | --- | --- |
| Phenotype | **Hazard ratio** | **p-value** | **95% CI** | **pval_fdr** | **Omics measure** |
| Female | | | | | |
| Hypertension | 1.081 | 0.24 | (0.95‚ 1.23) | 0.294 | Total T |
| Hypertension | 1.081 | 0.23 | (0.95‚ 1.23) | 0.294 | Bio T |
| Hypertension | 1.115 | 0.097 | (0.98‚ 1.27) | 0.241 | Free T |
| Hypertension | 1.102 | 0.34 | (0.9‚ 1.34) | 0.338 | E2 |
| Hypertension | 0.823 | 0.019 | (0.7‚ 0.97) | 0.095 | SHBG |
| Diabetes | 1.311 | 0.01 | (1.07‚ 1.61) | 0.0173 | Total T |
| Diabetes | 1.377 | 8.14E-05 | (1.17‚ 1.61) | 0.00041 | Bio T |
| Diabetes | 1.472 | 0.00038 | (1.19‚ 1.82) | 0.00095 | Free T |
| Diabetes | 1.174 | 0.102 | (0.97‚ 1.42) | 0.102 | E2 |
| Diabetes | 0.627 | 0.016 | (0.43‚ 0.92) | 0.0199 | SHBG |
| Male | | | | | |
| Hypertension | 1.034 | 0.5 | (0.94‚ 1.14) | 0.528 | Total T |
| Hypertension | 1.052 | 0.34 | (0.95‚ 1.17) | 0.528 | Bio T |
| Hypertension | 1.109 | 0.046 | (1‚ 1.23) | 0.114 | Free T |
| Hypertension | 1.12 | 0.0078 | (1.03‚ 1.22) | 0.0388 | E2 |
| Hypertension | 0.968 | 0.53 | (0.88‚ 1.07) | 0.528 | SHBG |
| Diabetes | 0.852 | 0.057 | (0.72‚ 1) | 0.0945 | Total T |
| Diabetes | 0.969 | 0.67 | (0.84‚ 1.12) | 0.668 | Bio T |
| Diabetes | 1.221 | 0.0054 | (1.06‚ 1.41) | 0.027 | Free T |
| Diabetes | 1.034 | 0.61 | (0.91‚ 1.18) | 0.668 | E2 |
| Diabetes | 0.814 | 0.013 | (0.69‚ 0.96) | 0.0318 | SHBG |

Total T: total testosterone; Bio T: bioavailable testosterone; Free T: free testosterone;

E2: estradiol; SHBG: sex hormone binding protein.

| Table S7. Association analysis for metabolite scores (MetS) of sex hormones and sleep phenotypes at exam 5 | | | | |
| --- | --- | --- | --- | --- |
| Phenotype | Coefficient | p-value | 95% CI | Omics measure |
| Female | | | | |
| log_AHI | 0.19 | 0.03 | (0.02, 0.35) | Mets. Total T |
| WHIIRS | -0.34 | 0.21 | (-0.89, 0.2) | Mets. Total T |
| ESS | -0.04 | 0.90 | (-0.61, 0.53) | Mets. Total T |
| Insomnia | 0.00 | 0.98 | (-0.31, 0.32) | Mets. Total T |
| OSA_mild_sev | 0.20 | 0.31 | (-0.18, 0.59) | Mets. Total T |
| OSA_mod_sev | 0.24 | 0.19 | (-0.11, 0.6) | Mets. Total T |
| log_AHI | 0.13 | 0.14 | (-0.04, 0.31) | Mets. Bio T |
| WHIIRS | 0.17 | 0.58 | (-0.43, 0.77) | Mets. Bio T |
| ESS | -0.09 | 0.77 | (-0.7, 0.52) | Mets. Bio T |
| Insomnia | -0.20 | 0.25 | (-0.56, 0.14) | Mets. Bio T |
| OSA_mild_sev | 0.21 | 0.31 | (-0.19, 0.63) | Mets. Bio T |
| OSA_mod_sev | 0.10 | 0.60 | (-0.26, 0.46) | Mets. Bio T |
| log_AHI | 0.15 | 0.10 | (-0.03, 0.32) | Mets. Free T |
| WHIIRS | 0.37 | 0.22 | (-0.23, 0.97) | Mets. Free T |
| ESS | 0.15 | 0.62 | (-0.46, 0.77) | Mets. Free T |
| Insomnia | -0.21 | 0.24 | (-0.57, 0.14) | Mets. Free T |
| OSA_mild_sev | 0.18 | 0.36 | (-0.21, 0.59) | Mets. Free T |
| OSA_mod_sev | 0.16 | 0.39 | (-0.2, 0.53) | Mets. Free T |
| log_AHI | 0.11 | 0.18 | (-0.05, 0.28) | Mets. E2 |
| WHIIRS | -0.21 | 0.47 | (-0.76, 0.35) | Mets. E2 |
| ESS | 0.08 | 0.78 | (-0.49, 0.65) | Mets. E2 |
| Insomnia | 0.01 | 0.96 | (-0.32, 0.34) | Mets. E2 |
| OSA_mild_sev | 0.10 | 0.62 | (-0.28, 0.48) | Mets. E2 |
| OSA_mod_sev | 0.22 | 0.21 | (-0.12, 0.56) | Mets. E2 |
| log_AHI | -0.49 | 0.17 | (-1.2, 0.22) | Mets. SHBG |
| WHIIRS | -1.40 | 0.25 | (-3.8, 1) | Mets. SHBG |
| ESS | -0.67 | 0.59 | (-3.14, 1.79) | Mets. SHBG |
| Insomnia | 0.92 | 0.21 | (-0.49, 2.37) | Mets. SHBG |
| OSA_mild_sev | -0.61 | 0.46 | (-2.22, 1) | Mets. SHBG |
| OSA_mod_sev | -0.41 | 0.58 | (-1.88, 1.03) | Mets. SHBG |
| Male | | | | |
| log_AHI | -0.09 | 0.06 | (-0.18, 0.003) | Mets. Total T |
| WHIIRS | -0.28 | 0.16 | (-0.67, 0.11) | Mets. Total T |
| ESS | 0.24 | 0.19 | (-0.12, 0.6) | Mets. Total T |
| Insomnia | 0.11 | 0.27 | (-0.08, 0.3) | Mets. Total T |
| OSA_mild_sev | -0.35 | 0.02 | (-0.66, -0.05) | Mets. Total T |
| OSA_mod_sev | -0.05 | 0.62 | (-0.25, 0.15) | Mets. Total T |
| log_AHI | 0.03 | 0.54 | (-0.06, 0.12) | Mets. Bio T |
| WHIIRS | -0.40 | 0.05 | (-0.79, -0.01) | Mets. Bio T |
| ESS | 0.02 | 0.90 | (-0.34, 0.39) | Mets. Bio T |
| Insomnia | 0.23 | 0.02 | (0.03, 0.42) | Mets. Bio T |
| OSA_mild_sev | 0.20 | 0.19 | (-0.11, 0.5) | Mets. Bio T |
| OSA_mod_sev | 0.06 | 0.55 | (-0.14, 0.27) | Mets. Bio T |
| log_AHI | 0.10 | 0.03 | (0.01, 0.2) | Mets. Free T |
| WHIIRS | -0.05 | 0.80 | (-0.46, 0.36) | Mets. Free T |
| ESS | -0.32 | 0.10 | (-0.7, 0.06) | Mets. Free T |
| Insomnia | 0.05 | 0.62 | (-0.15, 0.25) | Mets. Free T |
| OSA_mild_sev | 0.49 | 0.002 | (0.18, 0.81) | Mets. Free T |
| OSA_mod_sev | 0.05 | 0.66 | (-0.17, 0.26) | Mets. Free T |
| log_AHI | 0.04 | 0.35 | (-0.05, 0.13) | Mets. E2 |
| WHIIRS | -0.44 | 0.02 | (-0.82, -0.06) | Mets. E2 |
| ESS | 0.03 | 0.87 | (-0.32, 0.38) | Mets. E2 |
| Insomnia | 0.28 | 0.004 | (0.09, 0.47) | Mets. E2 |
| OSA_mild_sev | 0.18 | 0.22 | (-0.11, 0.47) | Mets. E2 |
| OSA_mod_sev | 0.22 | 0.03 | (0.02, 0.42) | Mets. E2 |
| log_AHI | -0.41 | 0.05 | (-0.8, -0.001) | Mets. SHBG |
| WHIIRS | 0.11 | 0.91 | (-1.69, 1.9) | Mets. SHBG |
| ESS | 1.51 | 0.7 | (-0.15, 3.18) | Mets. SHBG |
| Insomnia | -0.16 | 0.72 | (-1.05, 0.72) | Mets. SHBG |
| OSA_mild_sev | -2.11 | 0.002 | (-3.49, -0.76) | Mets. SHBG |
| OSA_mod_sev | -0.16 | 0.75 | (-1.09, 0.78) | Mets. SHBG |
| AHI: apnea hypopnea index; WHIRS: Women's Health Initiative Insomnia Rating Scale; ESS: Epworth Sleepiness Scale; OSA: obstructive sleep epnea. Total T: total testosterone; Bio T: bioavailable testosterone; Free T: free testosterone; E2: estradiol; SHBG: sex hormone binding protein. | | | | |

| Table S8. Association analysis for protein scores (ProtS) of sex hormones and sleep phenotypes at exam 5 | | | | |
| --- | --- | --- | --- | --- |
| Phenotype | **Coefficient** | **p-value** | **95% CI** | **Omics measure** |
| Female | | | | |
| log_AHI | 0.02 | 0.64 | (-0.06, 0.1) | Prots. Total T |
| WHIIRS | 0.22 | 0.15 | (-0.08, 0.52) | Prots. Total T |
| ESS | -0.06 | 0.68 | (-0.32, 0.21) | Prots. Total T |
| Insomnia | -0.14 | 0.05 | (-0.28, 0.003) | Prots. Total T |
| OSA_mild_sev | 0.08 | 0.32 | (-0.08, 0.25) | Prots. Total T |
| OSA_mod_sev | 0.14 | 0.08 | (-0.02, 0.3) | Prots. Total T |
| log_AHI | 0.18 | 1.67E-05 | (0.1, 0.27) | Prots. Bio T |
| WHIIRS | -0.13 | 0.41 | (-0.46, 0.19) | Prots. Bio T |
| ESS | -0.09 | 0.52 | (-0.38, 0.19) | Prots. Bio T |
| Insomnia | -0.02 | 0.74 | (-0.17, 0.12) | Prots. Bio T |
| OSA_mild_sev | 0.25 | 0.01 | (0.07, 0.43) | Prots. Bio T |
| OSA_mod_sev | 0.30 | 0.0005 | (0.13, 0.47) | Prots. Bio T |
| log_AHI | 0.18 | 1.27E-05 | (0.1, 0.27) | Prots. Free T |
| WHIIRS | -0.05 | 0.75 | (-0.37, 0.27) | Prots. Free T |
| ESS | 0.02 | 0.89 | (-0.26, 0.3) | Prots. Free T |
| Insomnia | -0.02 | 0.77 | (-0.17, 0.12) | Prots. Free T |
| OSA_mild_sev | 0.25 | 0.01 | (0.07, 0.43) | Prots. Free T |
| OSA_mod_sev | 0.27 | 0.00168 | (0.1, 0.43) | Prots. Free T |
| log_AHI | 0.00 | 0.99 | (-0.08, 0.08) | Prots. E2 |
| WHIIRS | -0.17 | 0.26 | (-0.46, 0.13) | Prots. E2 |
| ESS | 0.03 | 0.80 | (-0.23, 0.29) | Prots. E2 |
| Insomnia | 0.00 | 0.98 | (-0.14, 0.13) | Prots. E2 |
| OSA_mild_sev | 0.00 | 0.97 | (-0.17, 0.16) | Prots. E2 |
| OSA_mod_sev | 0.01 | 0.91 | (-0.14, 0.16) | Prots. E2 |
| log_AHI | -0.18 | 1.37E-05 | (-0.26, -0.1) | Prots. SHBG |
| WHIIRS | 0.05 | 0.75 | (-0.27, 0.37) | Prots. SHBG |
| ESS | -0.03 | 0.85 | (-0.31, 0.25) | Prots. SHBG |
| Insomnia | 0.02 | 0.78 | (-0.12, 0.16) | Prots. SHBG |
| OSA_mild_sev | -0.25 | 0.01 | (-0.4, -0.07) | Prots. SHBG |
| OSA_mod_sev | -0.26 | 0.0019 | (-0.43, -0.1) | Prots. SHBG |
| Male | | | | |
| log_AHI | -0.04 | 0.29 | (-0.12, 0.04) | Prots. Total T |
| WHIIRS | 0.26 | 0.11 | (-0.06, 0.58) | Prots. Total T |
| ESS | 0.28 | 0.08 | (-0.03, 0.58) | Prots. Total T |
| Insomnia | -0.15 | 0.08 | (-0.32, 0.02) | Prots. Total T |
| OSA_mild_sev | 0.02 | 0.88 | (-0.21, 0.25) | Prots. Total T |
| OSA_mod_sev | -0.05 | 0.57 | (-0.22, 0.12) | Prots. Total T |
| log_AHI | -0.004 | 0.91 | (-0.08, 0.07) | Prots. Bio T |
| WHIIRS | -0.09 | 0.57 | (-0.41, 0.22) | Prots. Bio T |
| ESS | -0.01 | 0.94 | (-0.31, 0.29) | Prots. Bio T |
| Insomnia | 0.05 | 0.56 | (-0.11, 0.21) | Prots. Bio T |
| OSA_mild_sev | 0.07 | 0.56 | (-0.16, 0.3) | Prots. Bio T |
| OSA_mod_sev | 0.01 | 0.87 | (-0.15, 0.18) | Prots. Bio T |
| log_AHI | 0.01 | 0.83 | (-0.07, 0.08) | Prots. Free T |
| WHIIRS | -0.40 | 0.01 | (-0.71, -0.08) | Prots. Free T |
| ESS | -0.19 | 0.20 | (-0.49, 0.1) | Prots. Free T |
| Insomnia | 0.22 | 0.01 | (-0.06, 0.39) | Prots. Free T |
| OSA_mild_sev | -0.02 | 0.84 | (-0.25, 0.2) | Prots. Free T |
| OSA_mod_sev | -0.01 | 0.89 | (-0.18, 0.16) | Prots. Free T |
| log_AHI | -0.0014 | 0.97 | (-0.07, 0.07) | Prots. E2 |
| WHIIRS | 0.06 | 0.67 | (-0.22, 0.35) | Prots. E2 |
| ESS | -0.03 | 0.82 | (-0.3, 0.24) | Prots. E2 |
| Insomnia | 0.0008 | 0.99 | (-0.15, 0.15) | Prots. E2 |
| OSA_mild_sev | 0.05 | 0.61 | (-0.16, 0.27) | Prots. E2 |
| OSA_mod_sev | 0.03 | 0.70 | (-0.12, 0.18) | Prots. E2 |
| log_AHI | -0.01 | 0.80 | (-0.08, 0.07) | Prots. SHBG |
| WHIIRS | 0.39 | 0.01 | (0.08, 0.7) | Prots. SHBG |
| ESS | 0.18 | 0.24 | (-0.12, 0.47) | Prots. SHBG |
| Insomnia | -0.22 | 0.01 | (-0.39, -0.06) | Prots. SHBG |
| OSA_mild_sev | 0.02 | 0.88 | (-0.21, 0.24) | Prots. SHBG |
| OSA_mod_sev | 0.01 | 0.90 | (-0.16, 0.18) | Prots. SHBG |
| AHI: apnea hypopnea index; WHIRS: Women's Health Initiative Insomnia Rating Scale; ESS: Epworth Sleepiness Scale; OSA: obstructive sleep epnea. Total T: total testosterone; Bio T: bioavailable testosterone; Free T: free testosterone;  E2: estradiol; SHBG: sex hormone binding protein. | | | | |

### Supplementary Figures

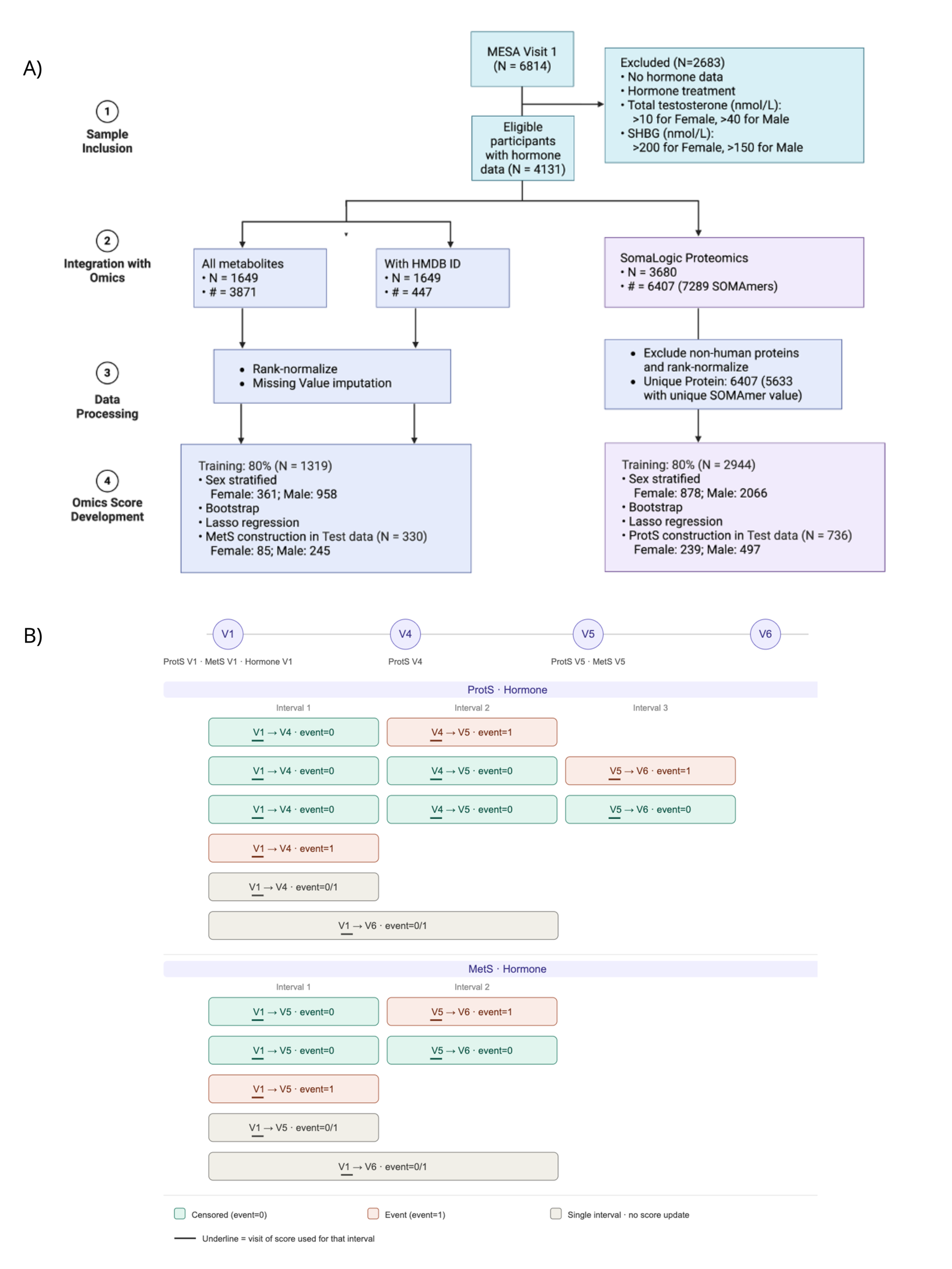

Figure S1. Data structure for counting process Cox model illustrating participant follow-up intervals across visits V1–V6. Protein (ProtS) and metabolite (MetS) scores updated at interval start (indicated by underline); green = censored interval, red = incident event, gray = single interval with no score update.

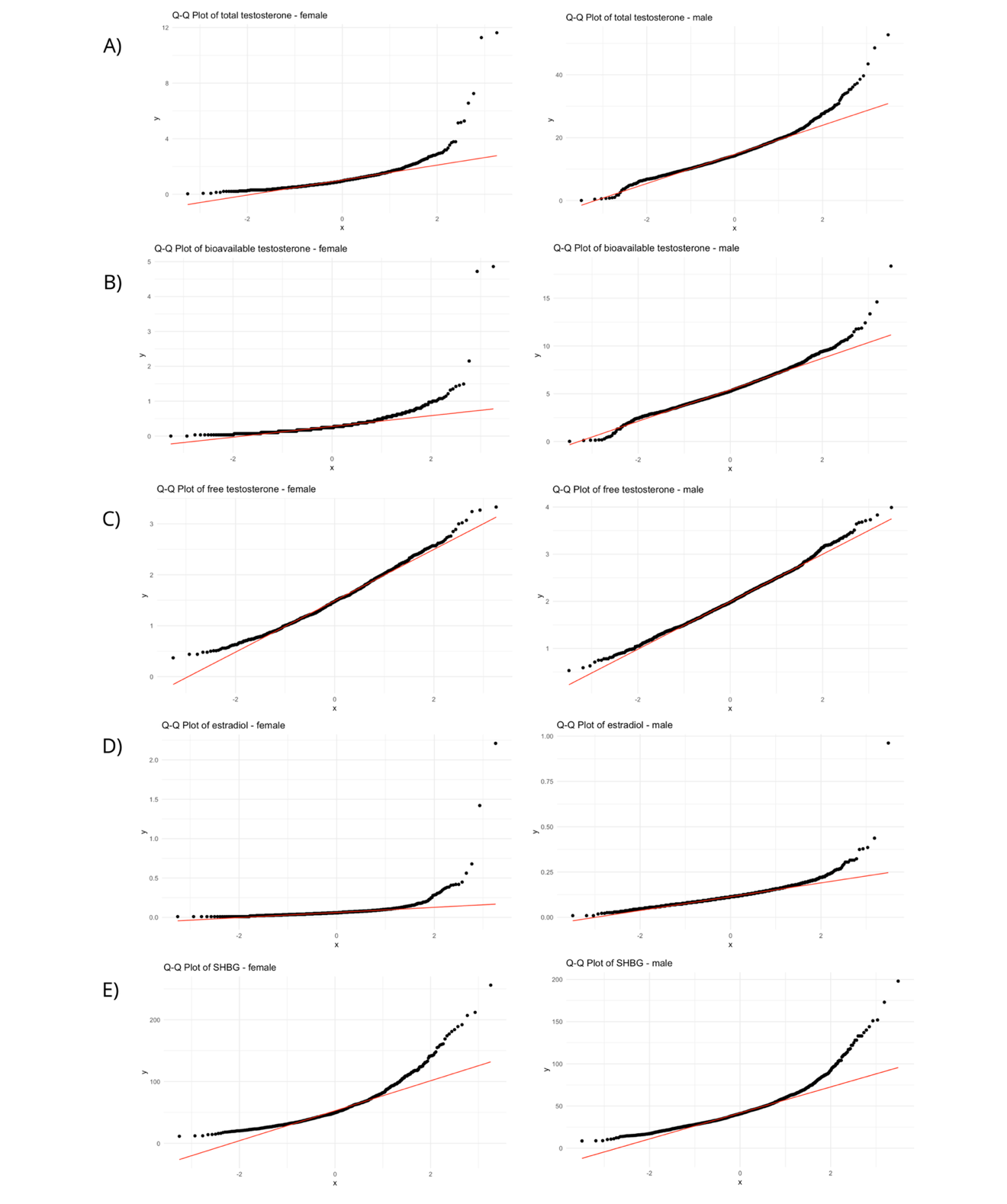

Figure S2. Quantile-quantile (QQ) plot for sex hormones in Female and Male MESA participants. A) Total testosterone; B) Bioavailable testosterone; C) Free testosterone; D) Estradiol; E) Sex hormone binding protein (SHBG).

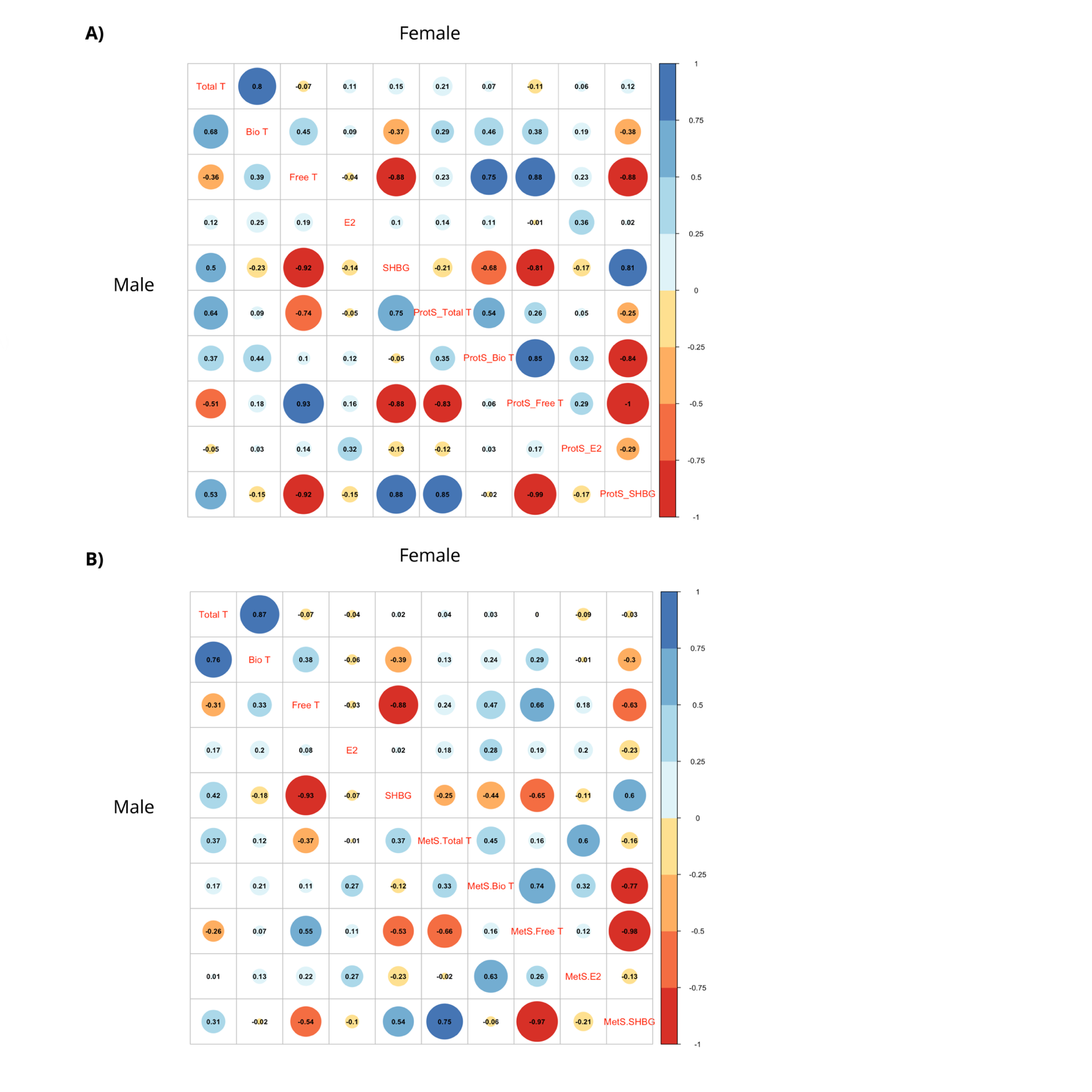

Figure S3. Correlation plot between sex hormones and A) protein (ProtS) and B) metabolite scores (MetS). Each of the two figures is split between females (top right triangle) and males (bottom left triangle). Total T: total testosterone; Bio T: bioavailable testosterone; Free T: free testosterone; E2: estradiol; SHBG: sex hormone binding protein. Var: variance

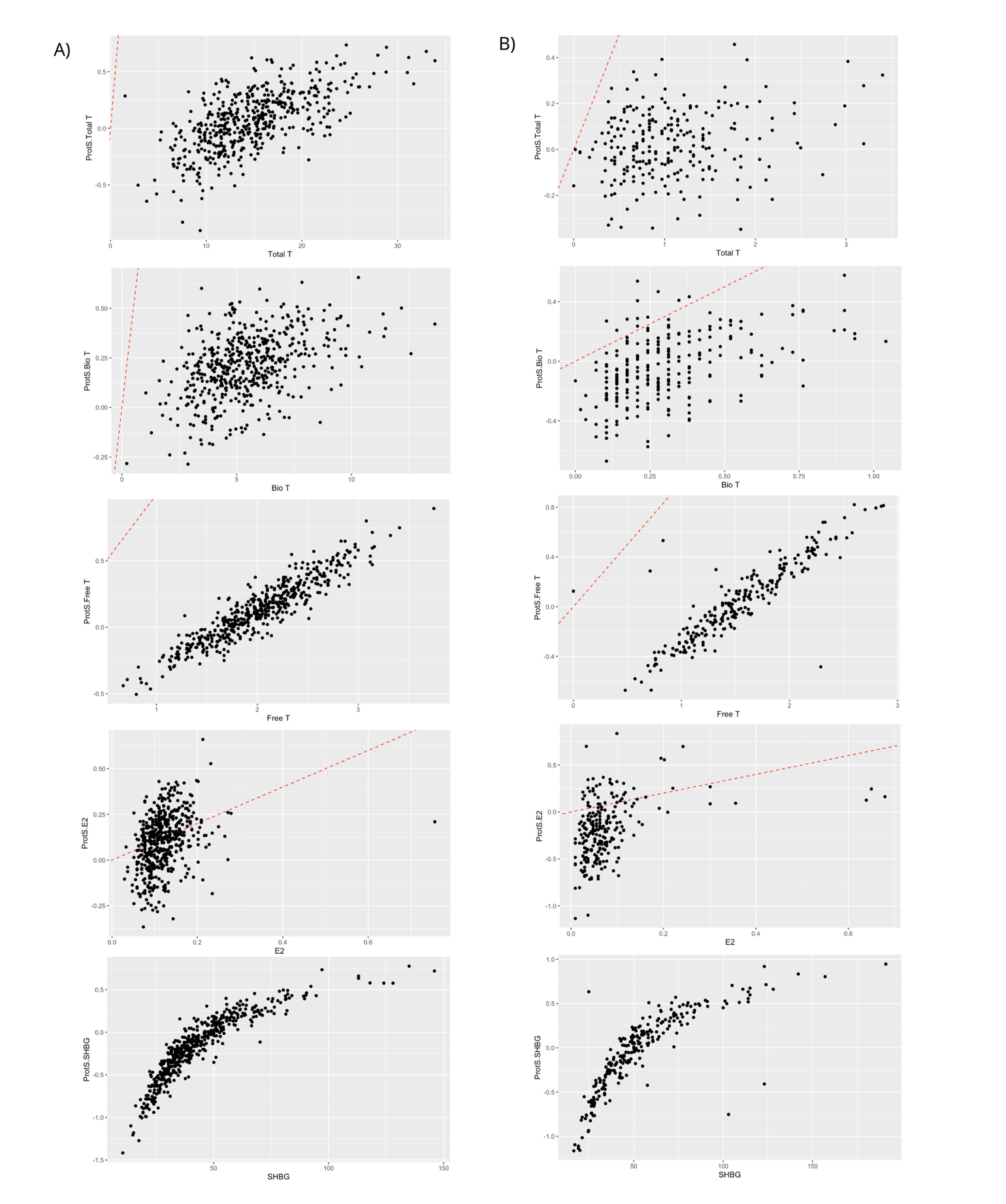

Figure S4. Scatterplot of protein scores (ProtS) against hormones. A) Male; B) Female

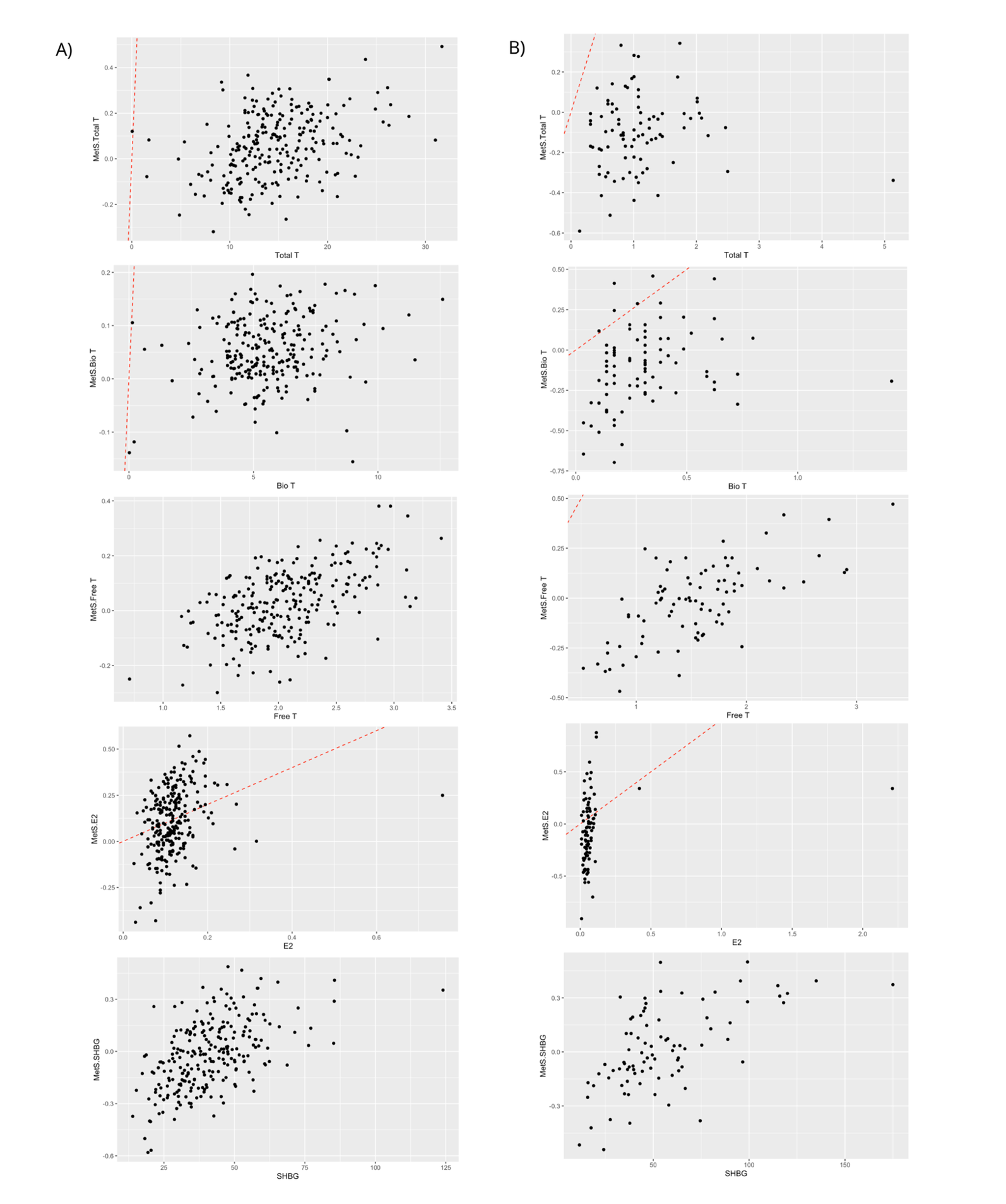

Figure S5. Scatterplot of metabolite scores (MetS) against hormones. A) Male; B) Female

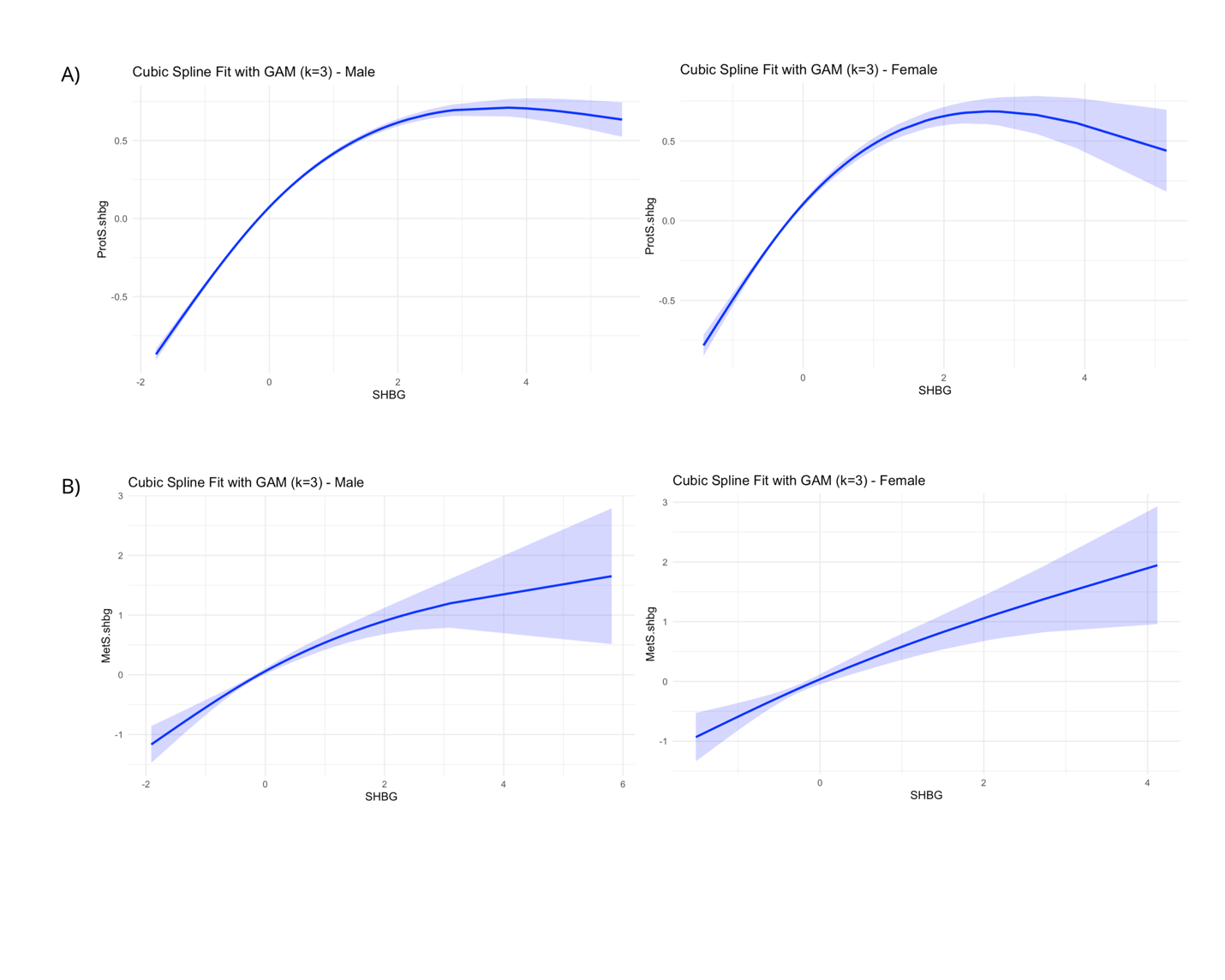

Figure S6. Cubic spline model fit plot for protein (ProtS) and metabolite scores (MetS) with sex hormone binding protein (SHBG). A) ProtS; B) MetS

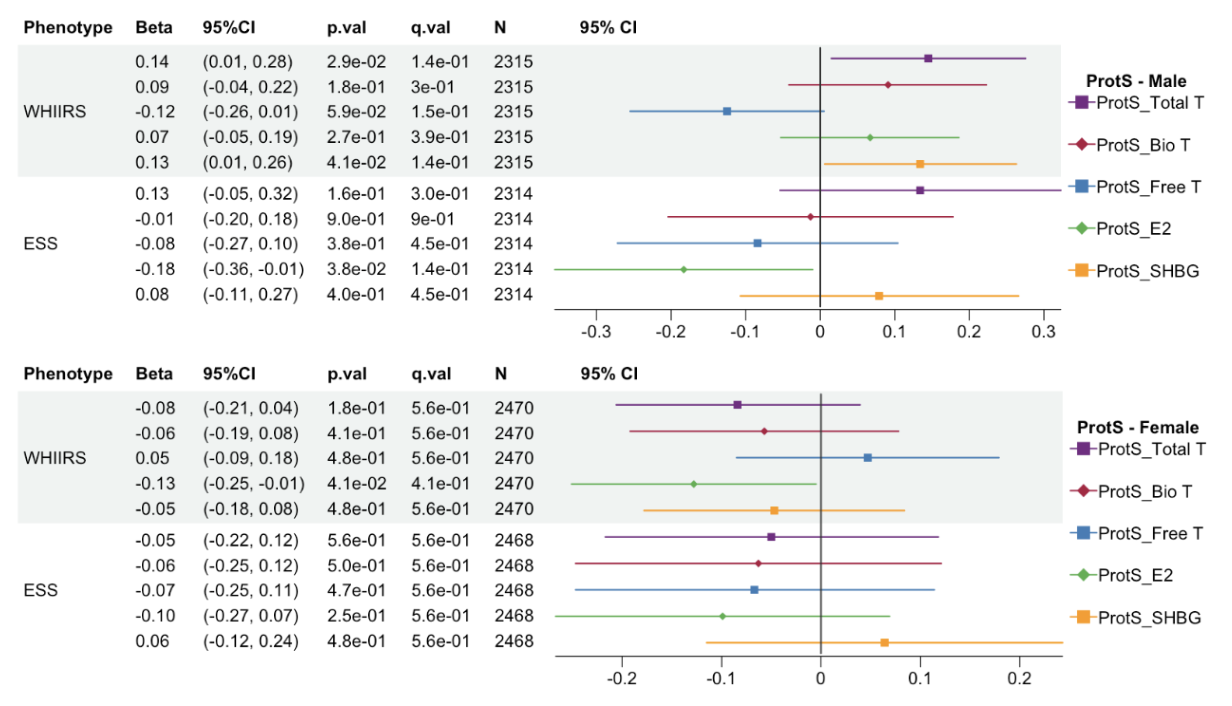

Figure S7. Forest plots of protein (ProtS) scores for sex hormones based associations across sleep phenotypes at exam 4, stratified by female and male participants. WHIRS: Women's Health Initiative Insomnia Rating Scale; ESS**:** Epworth Sleepiness Scale. Total T: total testosterone; Bio T: bioavailable testosterone; Free T: free testosterone; E2: estradiol; SHBG: sex hormone binding protein.
